# Association of age with clinical progression across plasma p-tau217 levels

**DOI:** 10.64898/2026.08.08.26359816

**Authors:** Ruyu Shi, Lamia T. Choity, Shayna T. Brodman, Xuemei Zeng, Marissa F. Farinas, Michel N. Nafash, Alexandra Gogola, Brian Lopresti, Dana L. Tudorascu, Sarah B. Berman, Robert A. Sweet, Victor L. Villemagne, Julia Kofler, C. Elizabeth Shaaban, Milos D. Ikonomovic, Tharick A. Pascoal, Ann D. Cohen, Oscar L. Lopez, Beth E. Snitz, M. Ilyas Kamboh, Thomas K. Karikari

## Abstract

**BACKGROUND:** Chronological age and plasma p⍰tau217 each predicts cognitive decline, but whether their prognostic associations interact is unclear. In this study, we examined their joint associations in a memory-clinic cohort.

**METHODS:** We included 3,741 participants from the Pittsburgh ADRC, with up to 28 years of follow-up (3.0 [IQR 2.0-6.0]). The primary outcome was increase in Clinical Dementia Rating global score (CDR-GS). Secondary outcomes included clinical-stage progression and longitudinal change in CDR Sum of Boxes. Plasma p-tau217 cut-off value was derived and externally validated in amyloid-beta-PET and autopsy sub-cohorts, respectively. Cox proportional hazards and linear mixed-effects models tested age-by-p-tau217 interactions while repeated cross-validation evaluated prognostic performance.

**RESULTS:** Age and plasma p-tau217 interacted in their associations with CDR-GS progression (χ(1)^2^=23.94; p=9.81×10^-7^). Comparing the oldest displayed age with the youngest reference age, the adjusted hazard ratio was 4.80 (95% CI 2.74-8.27) in the lowest vs. 1.05 (95% CI 0.69-1.47) in the highest p-tau217 quartile. Older age was associated with clinical progression at low-p-tau217 (HR=1.97; 95% CI 1.58-2.45) but not at high-p-tau217 (HR=1.10; 95% CI 0.95-1.26) concentrations; adjusted 5-year risk differences were 19.3 and 3.3 percentage points, respectively. Adding plasma p-tau217 improved 5-year discrimination most accurately among participants younger than 60 years (AUC 0.66-0.81).

**DISCUSSION:** Prognostic association between age and clinical progression varies by plasma p-tau217 concentration. Age stratifies risk at low plasma p-tau217 levels, whereas elevated p-tau217 identifies higher risk across age groups and attenuates the age-related gradient. These findings support further evaluation of age-contextualized plasma p-tau217 interpretation for prognosis and trial enrichment.

## 1 BACKGROUND

Alzheimer’s disease and related dementias (ADRD) account for a substantial proportion of the global burden of neurological disorders and are associated with major mortality, morbidity and economic costs.^1^ Older age remains the strongest epidemiological risk factor for cognitive decline and is routinely incorporated into clinical risk assessment, epidemiology models, and therapeutic trial design.^2,3^ However, individuals of similar chronological age often show marked heterogeneity in cognitive trajectories.^4,5^ This heterogeneity may reflect differences in Alzheimer’s disease (AD) pathology and clinical manifestation.^6,7^ As a result, chronological age alone has limited specificity for identifying individuals at highest risk for near-term cognitive decline.

Blood-based biomarkers offer a transformative opportunity to refine the interpretation of age-related risk. Plasma phosphorylated tau at amino acid threonine-217 (p-tau217) is closely associated with core AD pathology and correlates strongly with *in vivo* amyloid-β (Aβ) and tau positron-emission tomography (PET) abnormalities.^8–12^ Although plasma p-tau217 has shown robust performance for identifying amyloid-β pathology, its performance for predicting disease progression is more modest, likely reflecting the broader clinical and biological heterogeneity that shapes cognitive decline. This raises an important question that whether plasma p-tau217 simply adds prognostic information beyond age, or whether it fundamentally changes the prognostic meaning of age itself.

The distinction is clinically important because age may not carry the same weight across biomarker-defined groups. An older individual with low biomarker evidence of AD pathology may not have the high progression risk expected from age alone, whereas a younger individual with elevated plasma p-tau217 may have an accelerated risk profile that resembles, or even exceeds, that of older biomarker-positive individuals. Younger age may therefore provide a false sense of protection, particularly because early-onset AD is often associated with faster clinical decline.^13^ Accurate risk stratification across the age spectrum is therefore essential for identifying individuals who may warrant closer monitoring, earlier intervention, or enrollment in therapeutic trials.

Clarifying this relationship is furthermore essential for the clinical implementation and actionable interpretation of blood-based biomarkers. If age-related progression risk differs according to plasma p-tau217 status, then p-tau217 may support age-contextualized risk stratification rather than function only as an additive predictor in standard multivariable models. Such an approach could improve blood-based screening strategies, enrich therapeutic trials with participants at higher near-term risk of progression, and provide negative prognostic information for older biomarker-low individuals who might otherwise be considered high risk based on age alone.

Here, using a large, well-characterized, longitudinal, memory-clinic-based ADRC cohort spanning the Alzheimer’s disease continuum, we examined the joint association of chronological age and plasma p-tau217 with clinical progression. Specifically, we tested whether the prognostic association between age and progression risk is modified by plasma p-tau217 status, moving beyond incremental predictive metrics to characterize age-related risk gradients within biomarker-defined strata. We evaluated 3,741 participants followed for up to 28 years, with detailed cognitive assessments, plasma biomarker assessments, brain Aβ plaque imaging, and post-mortem neuropathological evaluation in relevant subgroups.

## 2. METHODS

### 2.1 Study population

Participants were enrolled through the University of Pittsburgh Alzheimer’s Disease Research Center (Pitt-ADRC) from 1993 through 2023. Pitt-ADRC is a National Institute on Aging-funded academic memory center. General eligibility required at least seven years of formal education, adequate visual and auditory acuity for neuropsychological testing, and the availability of a reliable study partner. Exclusion criteria included major neurological or psychiatric disorders unrelated to neurodegeneration, recent electroconvulsive therapy, current substance use disorders, cancer within the past five years (except non-melanoma skin cancer and in situ prostate cancer), or any significant medical condition that could interfere with neuropsychological assessment. Detailed descriptions of recruitment procedures and clinical assessments have been published previously.^14^ In the present study, we selected participants based on the availability of longitudinal clinical follow-up information including Clinical Dementia Rating global score (CDR-GS), as well as baseline plasma biomarker measurements. A subset of participants underwent [^11^C] Pittsburgh Compound B (PiB) positron emission tomography (PET) to assess cerebral Aβ plaque burden.^15–17^ Another subset donated their brains and underwent post-mortem neuropathological assessments following the National Alzheimer’s Coordinating Center 749 (NACC) data collection procedures (versions 1, 7, 9, 10, and 11).^18^ In the PET cohort, Aβ status was defined using Magnetic Resonance Imaging (MRI)-independent Centiloid (CL)-scaled indices of Aβ burden, with >20 CL as A+.^19^ In the autopsy cohort, Aβ status was defined neuropathologically using the CERAD neuritic plaque score^20^. These data were used in the corresponding discovery and validation analyses for defining plasma p-tau217 cutoff values. Detailed neuroimaging and neuropathological assessment procedures have been previously described.^15,16,21^ The study was approved by the University of Pittsburgh Institutional Review Board (MOD19110245023).

### 2.2 Assessment of cognitive status and clinical diagnosis

Details regarding the clinical diagnostic framework, including neuropsychological and psychiatric evaluation and brain MRI protocols, have been documented in a previous study.^22^ At baseline and during subsequent annual follow-ups, cognitive status was staged using the Clinical Dementia Rating (CDR). This clinician-rated instrument relies on structured interviews with both participants and informants to assess six functional domains: memory, orientation, judgement and problem solving, community affairs, home and hobbies, and personal care.^23^ Domain ratings reflect cognitive-related functional decline relative to the baseline, with the final global score (CDR-GS; ranging from 0 to 3) derived via the standard CDR algorithm, where higher score denotes more severe impairment.^23^ The CDR Sum of Boxes (CDR-SB), ranging from 0 to 18, was calculated by summing the six domain ratings and used as a continuous measure of cognitive and functional impairment, with higher scores indicating greater impairment.^24^ Simultaneously, clinical diagnoses were established through a multidisciplinary consensus process. A panel of neurologists, psychiatrists, and dementia specialists reviewed all clinical, laboratory, and MRI data against standardized criteria. Participants were then classified into four broad groups, cognitively unimpaired (CU), mild cognitive impairment (MCI), AD dementia, and non-AD dementia, where the latter were excluded from the current analyses.

### 2.3 Biomarker measures

Venous blood was collected and processed in accordance with previously established procedures.^25^ Samples were drawn into 10 mL K2-EDTA tubes (BD Biosciences, Cat. No. 366643) and centrifuged at 2,000 × g for 10 minutes at 4 °C within 2 hours of collection. Plasma was carefully separated, aliquoted, and stored at −80 °C until analysis. Buffy coat fractions were retained for *APOE* genotyping, which was performed as described in a previous publication.^26^

Plasma concentrations of p-tau217 was measured using Single molecule array (Simoa) assays on the HD-X platform (Quanterix, Billerica, MA, USA) as described by Zeng and colleagues.^27^ Briefly, plasma samples were thawed at room temperature and centrifuged at 4,000 × g for 10 minutes at 4°C before analysis to remove particulate matter. P-tau217 was measured using the ALZpath p-tau 217 V2 Assay (#104371). Quality-control samples at three concentration levels were included at the beginning and end of each assay run to assess reproducibility. The average within-plate coefficients of variation (CV) and between-plate CV were 8.1% and CV=6.9%, respectively.

### 2.4 Statistical analysis

All statistical analyses were performed in R version 4.5.1 (R Foundation for Statistical Computing, Vienna, Austria). Continuous variables are presented as medians with interquartile ranges (IQRs), and categorical variables as counts and percentages. Two-group continuous comparisons used Wilcoxon rank-sum tests. Comparisons across more than two groups used Kruskal-Wallis tests followed, when applicable, by Bonferroni-adjusted Dunn tests. Categorical comparisons used Pearson chi-square or Fisher exact tests, as appropriate.

The primary progression outcome was an increase in CDR-GS. Participants with baseline CDR-GS of 3 or without a follow-up CDR assessment were not eligible. The secondary progression outcome was clinical-diagnosis progression from cognitively unimpaired (CU) to mild cognitive decline (MCI) or Alzheimer’s disease (AD), or from MCI to AD dementia. Participants with baseline AD dementia, non-AD dementia, or no eligible follow-up diagnosis were excluded from this outcome. Participants without progression were censored at their last eligible assessment. Cox proportional hazards models were adjusted for sex, education, APOE ε4 carrier status, and self-reported race if not specified.

For the primary age-by-p-tau217 interaction analysis, age was modeled using a penalized spline with 3 degrees of freedom, whereas plasma p-tau217 was log-transformed and standardized. Age and p-tau217 values were winsorized at their 2nd and 98th percentiles to limit the influence of extreme observations, and the 2^nd^ percentile was the reference. Pointwise 95% confidence intervals (CIs) were estimated using 1,000 bootstrap resamples. Pointwise 95% CIs were obtained from 1,000 bootstrap resamples. Parallel analyses used clinical-stage progression and age groups younger than 70 years or 70 years or older.

The single- and dual-threshold p-tau217 cut-off values were derived using receiver operating characteristic (ROC) analyses in the non-autopsied Aβ-PET sub-cohort, restricted to Aβ-PET assessments within 2 years of blood collection. When more than one eligible scan was available, the closest scan was retained. The single cut-off value was determined by maximizing the Youden index. The dual-cutpoint approach defined low, intermediate, and high groups, with thresholds selected to provide ≥90% sensitivity and ≥90% specificity while limiting the intermediate group to no more than 20% of participants.^28–30^ PET-derived thresholds were applied without re-estimation in the autopsy sub-cohort. Exact binomial 95% CIs were reported for age-specific sensitivity and specificity. These derived thresholds were highly similar to those from a recent publication.^31^ Area under the curve (AUC), sensitivity, specificity, and correspondingly 95% confidence intervals (CIs) were reported.

For threshold-based progression analyses, plasma p-tau217 was classified as low (<0.546 pg/mL) or high (≥0.546 pg/mL) based on the data-driven approach described above, and age as younger than 70 years or 70 years or older. This age cutpoint was selected pragmatically as a clinically interpretable whole-decade threshold that approximates the reported 68-72-year inflection window for plasma p-tau217 and other neurodegenerative biomarkers.^32^ Unadjusted progression was summarized with Kaplan-Meier curves, log-rank tests, median progression-free time, incidence rates, and incidence rate ratios. Adjusted Cox models estimated age contrasts within each p-tau217 group and p-tau217 contrasts within each age group. Analyses were repeated for clinical-stage progression. Analogous CDR-GS models evaluated age contrasts within the low, intermediate, and high dual-threshold groups. Unadjusted cumulative risks were estimated at 2, 5, and 10 years. Covariate-standardized 5-year risks were estimated from an adjusted Cox model with an age-group-by-p-tau217-group interaction. Standardized risks were averaged over the covariate distribution of the analytic sample. Risk differences and 95% CIs were estimated with 1,000 bootstrap resamples.

Longitudinal CDR-SB trajectories were first analyzed over all available follow-up with linear mixed-effects models. Fixed effects included time, plasma p-tau217 group, age group, and all interactions. Participant-specific random intercepts and slopes were included when supported; random-intercept-only models were used after a singular fit. Models were fitted with maximum likelihood. Two-group p-tau217 models and the 4 joint age-p-tau217 groups were evaluated overall and within baseline CDR-GS strata of 0 and 0.5. A sensitivity analysis was restricted to the first 5 years. It compared linear time with piecewise-linear time with a knot at 2 years. Group-specific slopes for 0 to 2 years and greater than 2 to 5 years were obtained from fixed-effect contrasts.

Prediction analyses were restricted to participants with baseline CDR-GS of 0 or 0.5 and complete model covariates. The clinical Cox model included age, baseline CDR-GS, sex, self-reported race, years of education, and APOE ε4 carrier status. The augmented model also included log_2_-transformed p-tau217. Predictions used 20 repetitions of 5-fold cross-validation. Folds were stratified jointly by age group and progression status. In each training fold, models were fitted across all age groups. Averaged out-of-fold predictions were then evaluated within age groups younger than 60, 60 to 69, 70 to 79, and 80 years or older. Time-dependent AUCs at 2, 5, and 10 years used inverse probability of censoring weighting. AUC CIs and between-model differences used 1,000 bootstrap resamples. Prediction error was assessed with inverse probability of censoring-weighted Brier scores. Calibration compared mean predicted risks with Kaplan-Meier observed risks within tertiles for the group younger than 60 years and quintiles for the other age groups.

At the fixed Aβ-PET-derived threshold, prognostic positive predictive value (PPV), negative predictive value (NPV), sensitivity, and specificity were estimated at 2, 5, and 10 years within each age group. These estimates used censoring-adjusted Kaplan-Meier risks, with 1,000 bootstrap resamples for 95% CIs. The same threshold was evaluated for Aβ-PET classification in binary (<70 vs ≥70 years) and 10-year age groups. Threshold-positive prevalence was summarized in the same age groups. Exact binomial 95% CIs and Fisher exact tests for performance heterogeneity were used.

## 3. RESULTS

### 3.1 Participant characteristics and prognostic relevance of plasma p-tau217

A total of 3,741 participants with baseline CDR-GS measurements and plasma p-tau217 levels were included in the analysis (**Table 1**). Median age was 73.4 years (IQR 66.13-78.97 years), 59.9% were female, and 48.4% were APOE ε*4* carriers. Median plasma p-tau217 concentration was 0.80 pg/mL (IQR 0.38-1.39 pg/mL). Neuropathological diagnostic data were available for 407 (10.9%) participants. Aβ status was available for 605 (16.2%) participants, including 397 autopsied participants classified using neuritic plaque burden (338 positive and 59 negative) and 208 non-autopsied participants with Aβ PET imaging performed within two years of blood collection (106 positive and 102 negative) (**Table S1**). Plasma p-tau217 was substantially higher in Aβ-positive than Aβ-negative participants, with fold change (fc) of 3.13 in the PiB-PET sub-cohort (p=1.26×10^-37^) and 4.21 in the autopsy-sub-cohort (p=2.80×10^-20^).

**Table 1.**
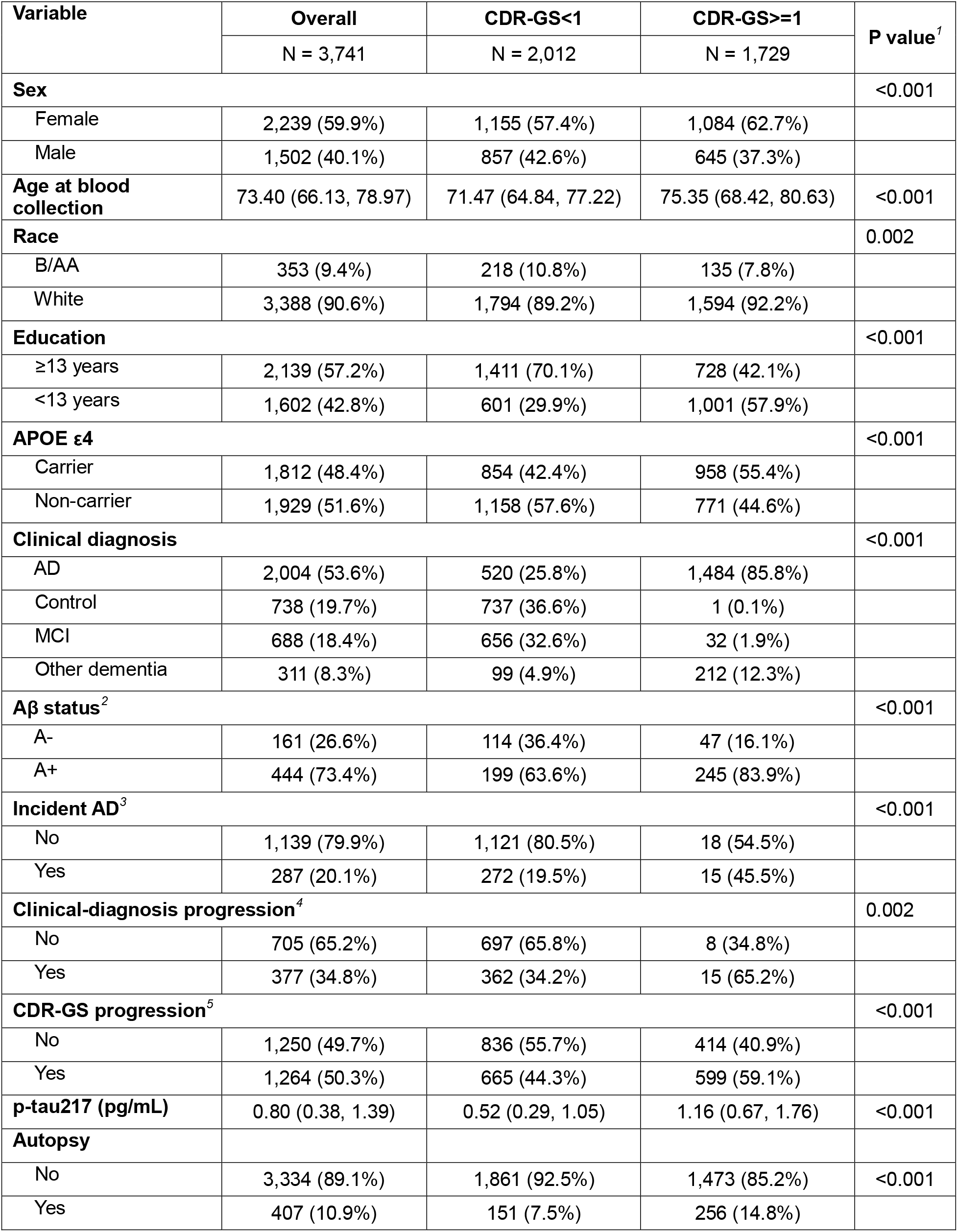
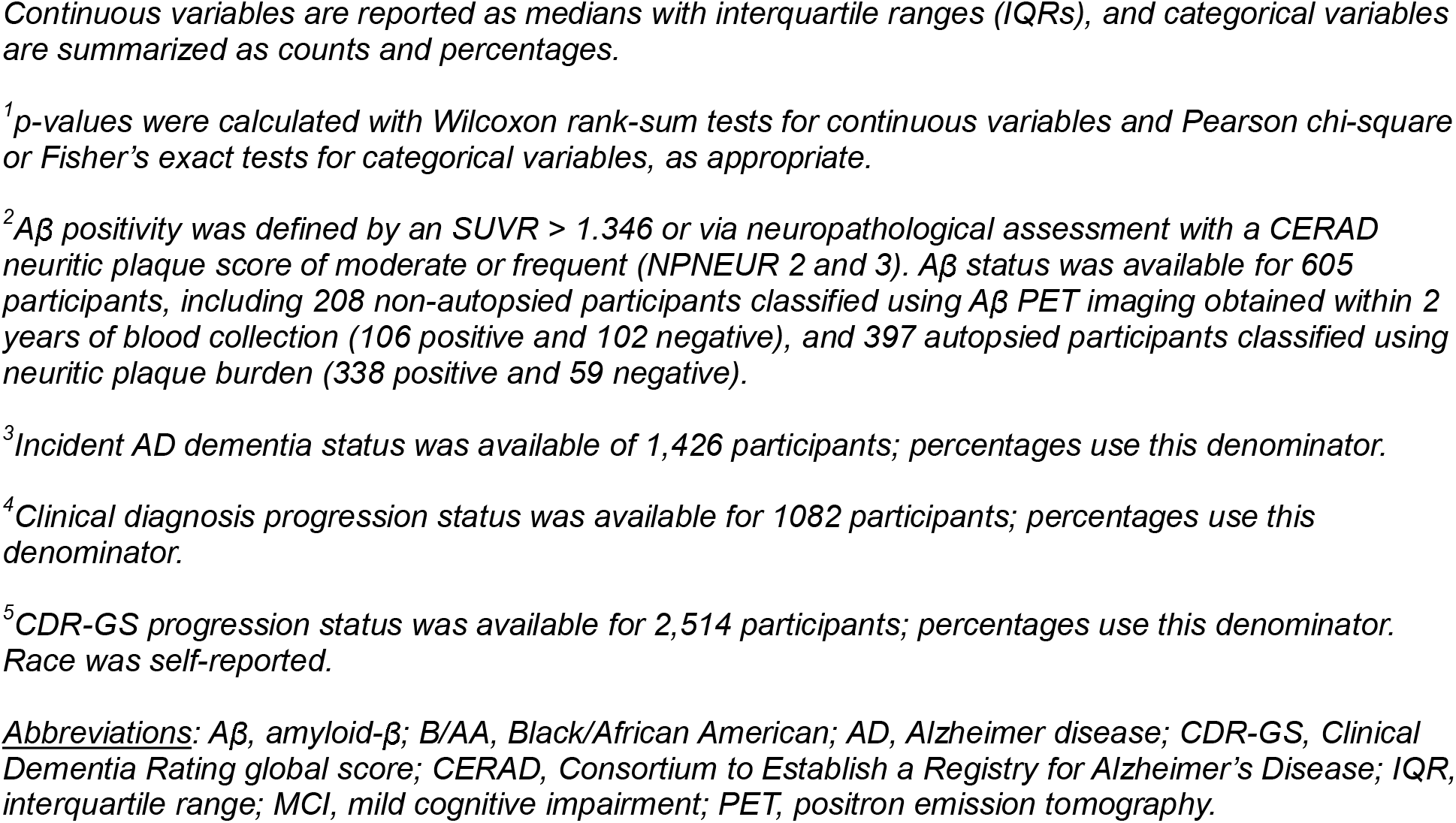
Baseline demographic and clinical characteristics by baseline CDR-GS group.

| Variable | Overall | CDR-GS<1 | CDR-GS>=1 | P value<sup>1</sup> |
| --- | --- | --- | --- | --- |
|  | N = 3,741 | N = 2,012 | N = 1,729 |  |
| **Sex** |  |  |  | <0.001 |
| Female | 2,239 (59.9%) | 1,155 (57.4%) | 1,084 (62.7%) |  |
| Male | 1,502 (40.1%) | 857 (42.6%) | 645 (37.3%) |  |
| **Age at blood collection** | 73.40 (66.13, 78.97) | 71.47 (64.84, 77.22) | 75.35 (68.42, 80.63) | <0.001 |
| **Race** |  |  |  | 0.002 |
| B/AA | 353 (9.4%) | 218 (10.8%) | 135 (7.8%) |  |
| White | 3,388 (90.6%) | 1,794 (89.2%) | 1,594 (92.2%) |  |
| **Education** |  |  |  | <0.001 |
| ≥13 years | 2,139 (57.2%) | 1,411 (70.1%) | 728 (42.1%) |  |
| <13 years | 1,602 (42.8%) | 601 (29.9%) | 1,001 (57.9%) |  |
| **APOE ε4** |  |  |  | <0.001 |
| Carrier | 1,812 (48.4%) | 854 (42.4%) | 958 (55.4%) |  |
| Non-carrier | 1,929 (51.6%) | 1,158 (57.6%) | 771 (44.6%) |  |
| **Clinical diagnosis** |  |  |  | <0.001 |
| AD | 2,004 (53.6%) | 520 (25.8%) | 1,484 (85.8%) |  |
| Control | 738 (19.7%) | 737 (36.6%) | 1 (0.1%) |  |
| MCI | 688 (18.4%) | 656 (32.6%) | 32 (1.9%) |  |
| Other dementia | 311 (8.3%) | 99 (4.9%) | 212 (12.3%) |  |
| **Aβ status<sup>2</sup>** |  |  |  | <0.001 |
| A- | 161 (26.6%) | 114 (36.4%) | 47 (16.1%) |  |
| A+ | 444 (73.4%) | 199 (63.6%) | 245 (83.9%) |  |
| **Incident AD<sup>3</sup>** |  |  |  | <0.001 |
| No | 1,139 (79.9%) | 1,121 (80.5%) | 18 (54.5%) |  |
| Yes | 287 (20.1%) | 272 (19.5%) | 15 (45.5%) |  |
| **Clinical-diagnosis progression<sup>4</sup>** |  |  |  | 0.002 |
| No | 705 (65.2%) | 697 (65.8%) | 8 (34.8%) |  |
| Yes | 377 (34.8%) | 362 (34.2%) | 15 (65.2%) |  |
| **CDR-GS progression<sup>5</sup>** |  |  |  | <0.001 |
| No | 1,250 (49.7%) | 836 (55.7%) | 414 (40.9%) |  |
| Yes | 1,264 (50.3%) | 665 (44.3%) | 599 (59.1%) |  |
| **p-tau217 (pg/mL)** | 0.80 (0.38, 1.39) | 0.52 (0.29, 1.05) | 1.16 (0.67, 1.76) | <0.001 |
| **Autopsy** |  |  |  |  |
| No | 3,334 (89.1%) | 1,861 (92.5%) | 1,473 (85.2%) | <0.001 |
| Yes | 407 (10.9%) | 151 (7.5%) | 256 (14.8%) |  |

Continuous variables are reported as medians with interquartile ranges (IQRs), and categorical variables are summarized as counts and percentages.
<sup>1</sup>p-values were calculated with Wilcoxon rank-sum tests for continuous variables and Pearson chi-square or Fisher's exact tests for categorical variables, as appropriate.
<sup>2</sup>Aβ positivity was defined by an SUVR > 1.346 or via neuropathological assessment with a CERAD neuritic plaque score of moderate or frequent (NPNEUR 2 and 3). Aβ status was available for 605 participants, including 208 non-autopsied participants classified using Aβ PET imaging obtained within 2 years of blood collection (106 positive and 102 negative), and 397 autopsied participants classified using neuritic plaque burden (338 positive and 59 negative).
<sup>3</sup>Incident AD dementia status was available of 1,426 participants; percentages use this denominator.
<sup>4</sup>Clinical diagnosis progression status was available for 1082 participants; percentages use this denominator.
<sup>5</sup>CDR-GS progression status was available for 2,514 participants; percentages use this denominator. Race was self-reported.
Abbreviations: Aβ , amyloid-β ; B/AA, Black/African American; AD, Alzheimer disease; CDR-GS, Clinical Dementia Rating global score; CERAD, Consortium to Establish a Registry for Alzheimer's Disease; IQR, interquartile range; MCI, mild cognitive impairment; PET, positron emission tomography.

Participants with baseline CDR-GS of 1 or higher were older than those with better cognitive functions (median 75.4 vs. 71.5 years), had less education (42.1% vs. 72.9%), and were more likely to be APOE ε4 carriers (958 of 1,729 [55.4%] vs 854 of 2,012 [42.4%]). The distribution of clinical diagnosis across CDR-GS categories is shown in **Figure S1A**. Baseline plasma p-tau217 showed stepwise increases across the spectrum of clinical impairment, from CDR-GS=0 through CDR-GS=3 (**Figure 1A**). Similar monotonic increase was observed in the clinical diagnosis group as well (**Figure S1B**). In adjusted-Cox proportional hazards models, higher baseline plasma p-tau217 was strongly associated with faster CDR-GS progression. Adjusted hazards increased monotonically across the observed biomarker range, with HRs of 1.56 (95%CI 1.44 to 1.71), 3.15 (95%CI 2.63 to 3.80), 5.06 (95%CI 4.14 to 6.27), and 10.47 (95%CI 7.04 to 15.97) at each p-tau217 quartile respectively (**Figure 1B**). A similar increase was observed for clinical diagnostic groups (**Figure S1C**).

**Figure 1.**
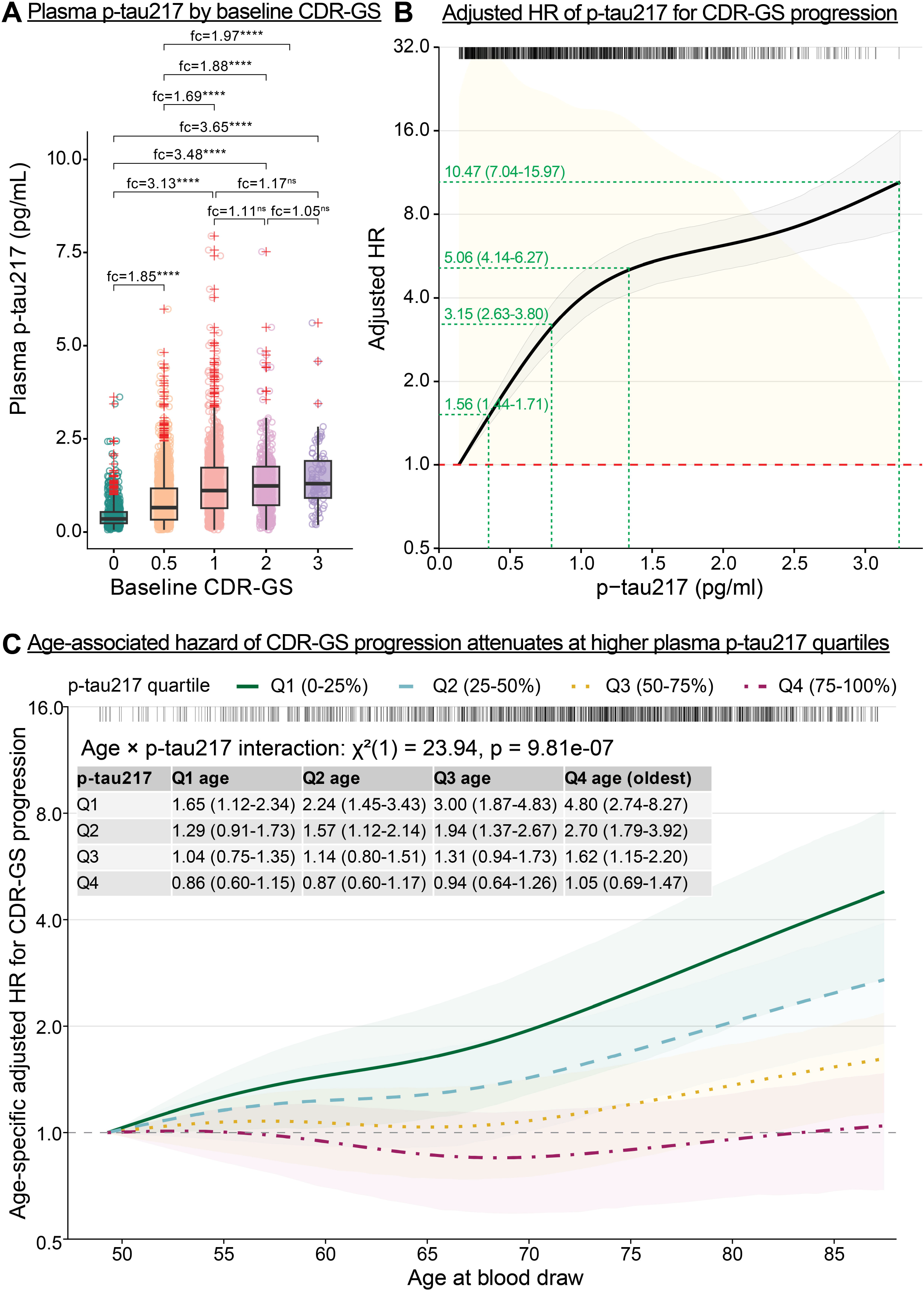
Plasma p-tau217 is associated with clinical severity and CDR-GS progression, with age-related risk attenuating at higher p-tau217 levels. A, Plasma p-tau217 distributions by baseline CDR-GS. Boxes represent the interquartile ranges (IQRs), and center lines indicate median. Group differences were assessed using the Kruskal-Wallis test followed by Dunn’s post hoc tests with false discovery rate (FDR) adjustment. Values above brackets indicate the fold change (fc) and statistical significance (*p < 0.05, **p < 0.01, ***p < 0.001, ****p < 0.0001). B, Adjusted HR of p-tau217 for CDR-GS progression. Dose-response curve showing the adjusted hazard ratio (HR) for progression to a higher CDR-GS according to p-tau217 levels (pg/mL). The solid black line indicates the estimated HR, and the shaded area indicates the 95% confidence interval (CI). The dashed red line marks an HR of 1.0. Risk increases with higher p-tau217 concentrations. C, Age-dependent hazard of CDR-GS progression by plasma p-tau217 quartile. Plot showing the adjusted HR for CDR-GS progression as a function of age at blood draw, stratified by p-tau217 quartiles (Q1-Q4). The association between age and progression risk is steeper in the lowest p-tau217 quartile (Q1, green solid line) than in the higher quartiles. The inset table provides specific HRs and 95% CIs for age groups across p-tau217 quartiles. <u>Abbreviations</u>: CDR-GS, Clinical Dementia Rating Global Score.

### 3.2 Age-by-p-tau217 interaction and CDR-GS progression

Older age was associated with faster CDR-GS progression (**Figure S1D**). This association differed substantially by plasma p-tau217 level (age-by-p-tau217 interaction likelihood-ratio χ^2^(1)=23.94, p=9.81×10^-7^) (**Figure 1C**). At the oldest displayed ages, relative to the youngest reference age, the adjusted HR for CDR-GS progression was 4.80 (95%CI 2.74 to 8.27) in the lowest p-tau217 quartile. The corresponding HRs were 2.70 (95% CI, 1.79-3.92), 1.62 (95% CI, 1.15-2.20), and 1.05 (95% CI, 0.69-1.47) in the second, third, and highest p-tau217 quartiles. The attenuation was also evident when age was dichotomized at 70 years and when clinical-stage progression was used (**Figure S1E-F**).

### 3.3 A**β** PET-derived plasma p-tau217 threshold and age-stratified progression

The Aβ-PET-derived single threshold for plasma p-tau217 was 0.546 pg/mL. In the 208-participant Aβ-PET sub-cohort for cutpoint estimation, sensitivity was 93.3% and specificity was 83.0%. In the displayed autopsy validation confusion matrix, sensitivity was 90.6% and specificity was 72.7% (**Figure S2A-C**). Aβ-PET classification performance was similar in the 2 broad age groups. Sensitivity was 96.1% (95% CI, 86.5%-99.5%) below age 70 years and 96.4% (95% CI 87.5% to 99.6%) at age 70 years or older. Specificity was 85.3% (95% CI 74.6% to 92.7%) and 79.4% (95% CI 62.1% to 91.3%), respectively (**Figure S2D**; **Table S2**). There was no evidence of heterogeneity by broad age group for sensitivity (p=1.00) or specificity (p=0.57). Estimates across 10-year age groups were also similar, although specificity was imprecise among participants aged 80 years or older (55.6%; 95% CI 21.2% to 86.3%; n=17). The prevalence of plasma p-tau217 at or above 0.546 pg/mL increased from 48.6% below age 70 years to 71.6% at age 70 years or older. Across 10-year groups, prevalence ranged from 45.7% below age 60 years to 78.0% at age 80 years or older (**Figure S1G-H**). Results from the dual-threshold approach (cutpoints: 0.444 and 0.690 pg/mL; intermediate group: 18.3%; sensitivity: 97.7%, specificity: 91.6% after excluding the intermediate group) are shown in **Figure S2E-G** and participant characteristics across the 4 joint groups are presented in **Table S3**. The low- and high-p-tau217 groups showed patterns consistent with those obtained using the single-cutoff approach, whereas the intermediate group more closely resembled the high-p-tau217 group.

The primary CDR-GS progression analysis included 2,514 participants, of whom 1,264 progressed. Incidence rates per 100 person-years were 5.04 and 10.43 in the low p-tau217 younger and older groups and 21.15 and 24.14 in the high p-tau217 groups (**Table S4A**). Unadjusted Kaplan-Meier curves differed across the 4 groups defined with age 70 and fixed p-tau217 of 0.546 pg/mL (log-rank p=1.06×10^-91^). Median progression-free times were 14.9, 6.2, 2.9, and 3.0 years, respectively (**Figure 2**). In adjusted models, older (≥70 years) vs younger age was associated with progression in the low p-tau217 group (HR=1.97, 95%CI 1.58 to 2.45) but not with the high p-tau217 group (HR=1.10, 95%CI 0.95 to 1.26). High vs low p-tau217 was associated with progression in both age groups, with a larger relative association among younger participants (HR=3.88, 95% CI 3.09 to 4.86) than among older participants (HR=2.51, 95% CI 2.12 to 2.97) (**Table S4B**).

**Figure 2.**
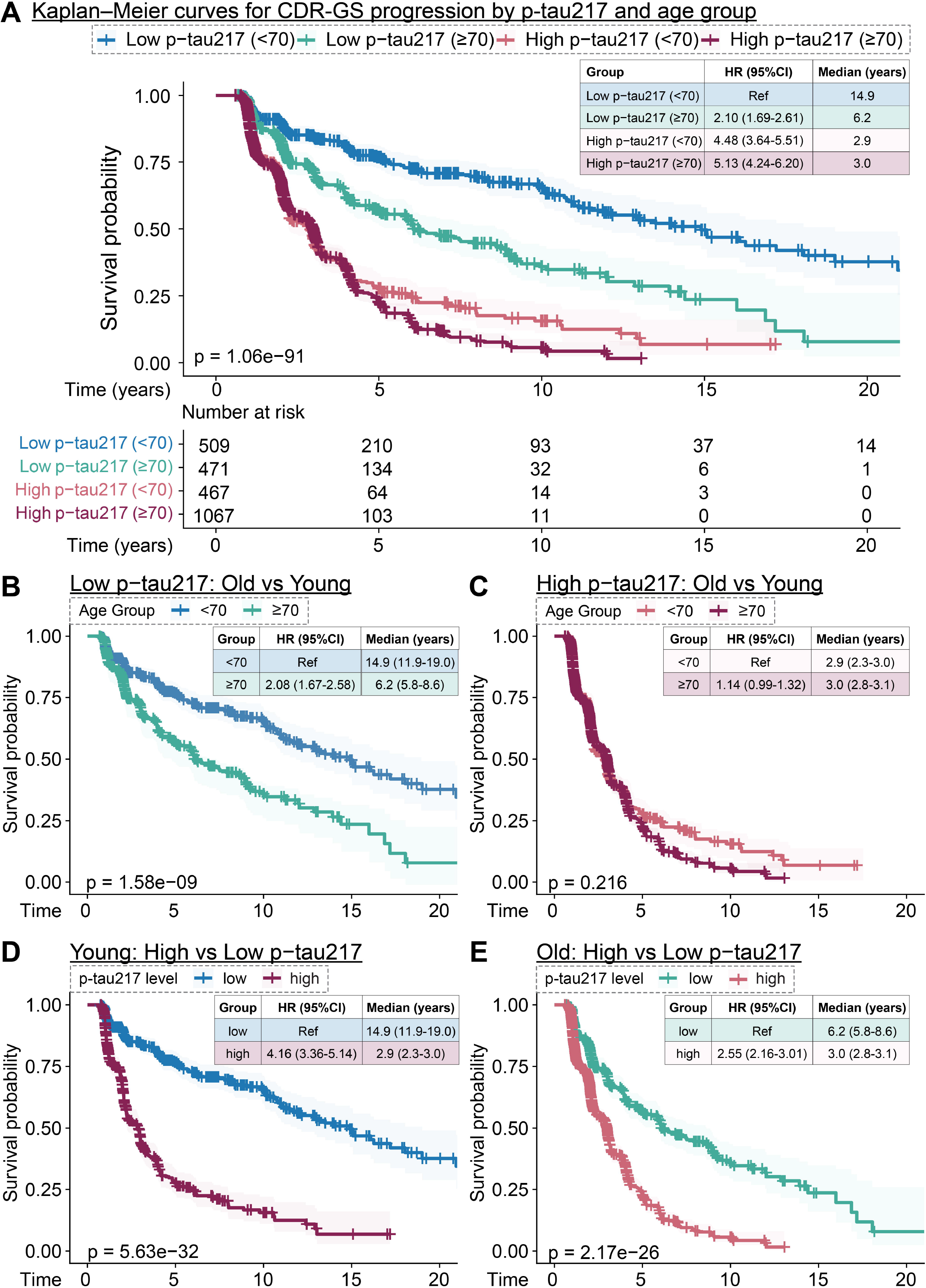
CDR-GS progression by PET-derived plasma p-tau217 threshold and age group. **A**, Kaplan-Meier curves for the 4 groups defined by age (<70 or ≥70 years) and the fixed p-tau217 threshold (low, <0.546 pg/mL; high, ≥0.546 pg/mL). **B-C**, Age comparisons within the low- and high-p-tau217 groups. **D-E**, P-tau217 comparisons within the younger and older groups. Annotated p-values correspond to comparisons of progression-free survival between groups. The embedded table reports unadjusted HRs, 95% CIs, and median progression-free survival for each group. <u>Abbreviations</u>: CDR-GS, Clinical Dementia Rating Global Score; HR, hazard ratio; CI, confidence interval.

For clinical progression, 377 of 1,082 participants progressed. Incidence rates per 100 person-years ranged from 2.37 in the low p-tau217 younger group to 18.05 in the high p-tau217 older group (**Table S5A**). The age association was stronger with low p-tau217 group (adjusted HR=2.93, 95% CI 2.08 to 4.12) than within the high p-tau217 group (adjusted HR=1.93, 95% CI, 1.43 to 2.61) (**Table S5B**).

Covariate-standardized 5-year risks showed the same pattern on an absolute scale (**Figure 3**; **Table S6**). With low p-tau217, estimated risk was 25.7% (95% CI 21.8% to 29.7%) for participants younger than 70 years and 45.0% (95% CI 40.6% to 49.5%) for those aged 70 years or older. The adjusted age-group risk difference was 19.3 percentage points (95% CI 13.5 to 25.0 percentage points). With high p-tau217, estimated risk was 71.5% (95% CI 66.5% to 76.0%) and 74.8% (95% CI 71.7% to 77.5%), respectively, resulting in an adjusted risk difference of 3.3 percentage points (95% CI −1.4 to 8.3 percentage points). In unadjusted analyses, the age-group difference within low p-tau217 widened from 7.1 percentage points at 2 years to 29.4 percentage points at 10 years. Differences within high p-tau217 remained smaller (**Figure S3A).** Baseline CDR-GS and diagnosis distributions differed across the 4 joint groups (**Figure S3B-C**).

**Figure 3.**
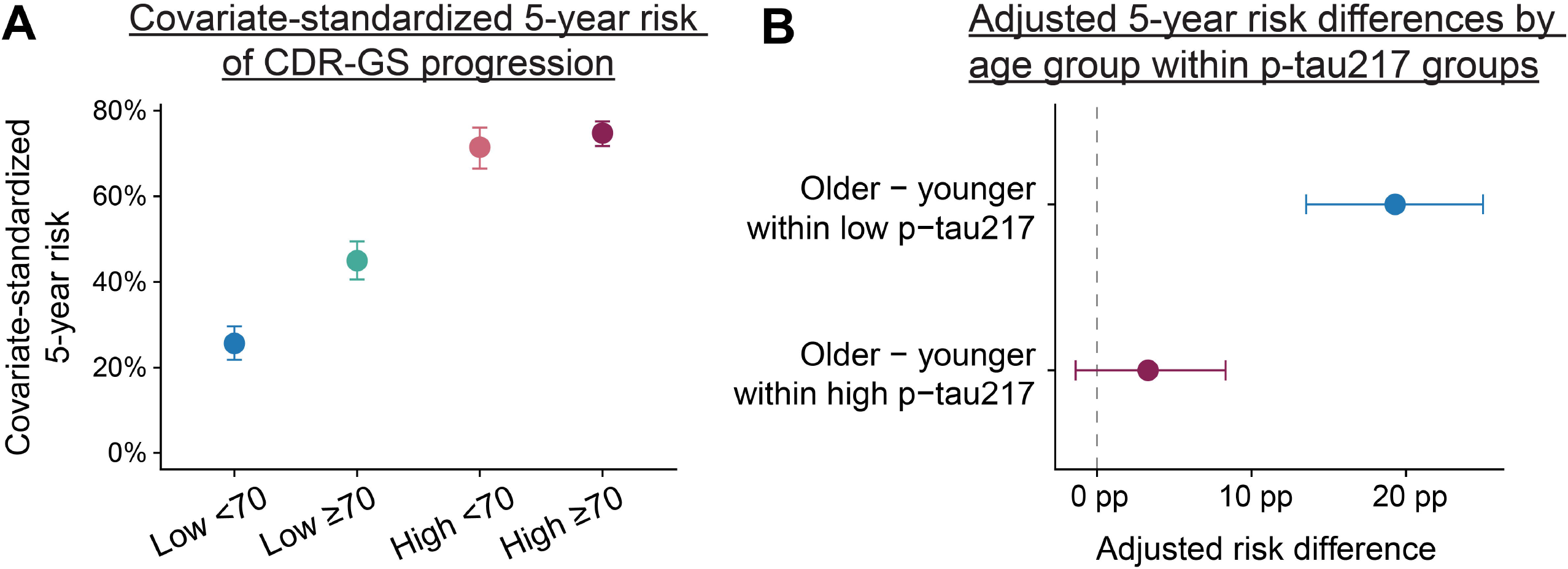
Covariate-standardized 5-year risk of CDR-GS progression. **A**, Covariate-standardized 5-year risks from a Cox model with an age-group-by-p-tau217-group interaction, adjusted for sex, education, APOE ε4, and self-reported race. Points are standardized risks and error bars are 95% bootstrap CIs. **B**, Adjusted risk differences for older (≥70 years) vs younger (<70 years) participants within each p-tau217 group. Low and high p-tau217 were defined by the PET-derived threshold of 0.546 pg/mL. <u>Abbreviations</u>: CDR-GS, Clinical Dementia Rating Global Score; CI, confidence interval; PET, positron emission tomography; pp, percentage points.

### 3.4 Longitudinal CDR-SB progression

The longitudinal analysis included 3,270 participants and 11,402 CDR-SB observations (median follow-up, 2.03 years; maximum, 28.83 years). In 2-group models, high vs low p-tau217 was associated with a 0.95-point-per-year greater CDR-SB increase overall, a 0.26-point greater increase among participants with baseline CDR-GS 0, and a 0.83-point greater increase among those with baseline CDR-GS 0.5 (all p<0.001) (**Figure 4A**). In the joint model, high p-tau217 was associated with faster worsening in the younger reference group (time-by-p-tau217 β=1.09; p=1.98×10^-47^), and older age was associated with faster worsening in the low-p-tau217 reference group (time-by-age β=0.24; p=6.59×10^-4^). The time-by-age-by-p-tau217 interaction was −0.29 (p=0.003), indicating greater age-related slope separation when p-tau217 was low (**Figure 4B**). Estimated annual increases were 0.28 (95% CI 0.19 to 0.38), 0.52 (95% CI 0.42 to 0.63), 1.37 (95% CI 1.27 to 1.48), and 1.32 (95% CI 1.25 to 1.39) points in the low-p-tau217-younger, low-p-tau217-older, high-p-tau217-younger, and high-p-tau217-older groups. In baseline CDR-GS-specific analyses, the 3-way estimate was positive for CDR-GS 0 (β=0.15; p=2.85×10^-8^) but was not significant for CDR-GS 0.5 (β = −0.24; p=0.065) (**Figure 4B**).

**Figure 4.**
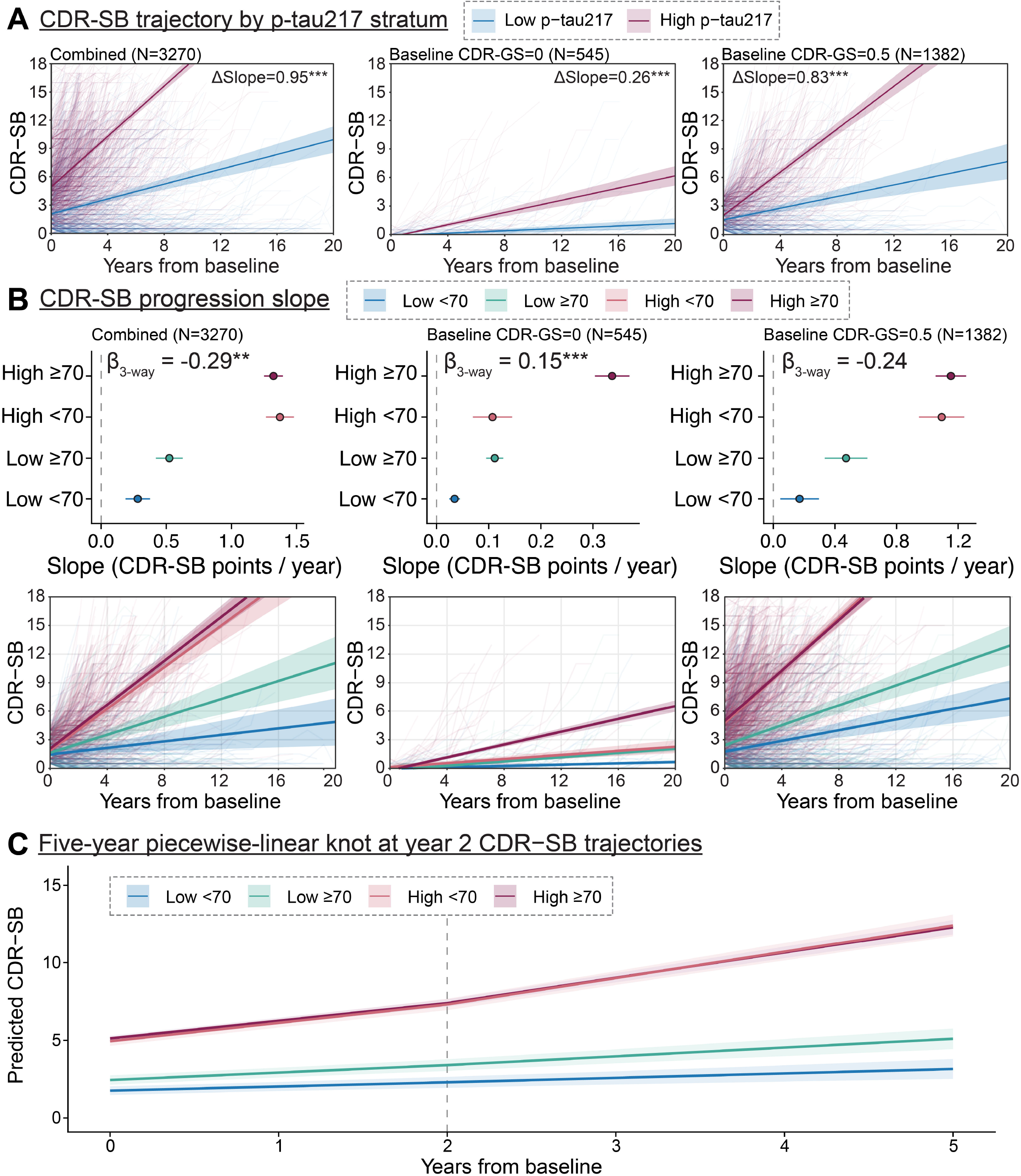
Longitudinal CDR-SB trajectories by plasma p-tau217 and age. **A**, Participant-level CDR-SB trajectories and fitted 2-group trajectories for low vs high p-tau217 overall and within baseline CDR-GS strata of 0 and 0.5. ΔSlope is the difference in annual CDR-SB change for high vs low p-tau217. **B**, Estimated annual slopes and fitted trajectories for the 4 joint age-p-tau217 groups. Forest plots show slope estimates with 95% CIs; β_3-way_ is the time-by-age-by-p-tau217 coefficient. Full-follow-up plots are displayed through 20 years. **C**, Predicted CDR-SB trajectories during the first 5 years from the piecewise-linear model with a knot at year 2. Bands show 95% CIs. Low and high p-tau217 were defined by 0.546 pg/mL; younger and older age by 70 years. **p<0.01; ***p<0.001. Abbreviations: CDR-GS, Clinical Dementia Rating Global Score; CDR-SB, Clinical Dementia Rating Sum of Boxes; CI, confidence interval.

In the sensitivity analysis restricted to 5 years, a piecewise-linear model with a knot at 2 years fit better than a constant linear-time model (likelihood-ratio χ^2^(4)=121.23; p=2.92×10^-25^). With low p-tau217, annual CDR-SB increases were 0.27 vs 0.48 points during years 0 to 2 and 0.29 vs 0.56 points during years greater than 2 to 5 for younger vs older participants, respectively. With high p-tau217, corresponding slopes were 1.18 vs 1.14 points and 1.70 vs 1.63 points. The time-by-age-by-p-tau217 interaction remained present in both intervals (p=0.032 and p=0.016, respectively) (**Figure 4C**; **Table S7**).

### 3.5 Age-stratified prediction performance of plasma p-tau217

Prediction analyses included 1,501 participants with baseline CDR-GS of 0 or 0.5, of whom 665 experienced CDR-GS progression. Adding log_2_-transformed p-tau217 improved 5-year time-dependent AUC most clearly in younger participants (**Figure S4A**; **Table S8A**). AUC increased from 0.66 to 0.81 below age 60 years (difference=0.15; 95% CI 0.07 to 0.24) and from 0.75 to 0.80 at ages 60 to 69 years (difference=0.06; 95% CI 0.02 to 0.10). Differences were smaller at ages 70 to 79 years (0.75 to 0.77; difference=0.03; 95% CI, −0.02 to 0.07) and 80 years or older (0.73 to 0.79; difference=0.05; 95% CI −0.04 to 0.15). At 2 and 10 years, statistically supported improvements were most consistent below age 60 years; the 10-year improvement was also supported at ages 70 to 79 years. Brier scores were lower with the augmented model at every age and time horizon. The largest 5-year reduction was below age 60 years (0.206 to 0.151) (**Table S8B**). Calibration varied across age and risk strata, especially in smaller groups (**Figure S4B**; **Table S8C**).

At the fixed threshold, 5-year NPV decreased from 86.4% (95% CI, 79.4%-93.0%) below age 60 years to 55.8% (95% CI, 42.2%-70.2%) at age 80 years or older. The corresponding PPVs were 64.3% (95% CI, 51.3%-78.0%) and 74.0% (95% CI, 64.4%-83.9%) (**Figure S4C**; **Table S8D**). Across age groups, 5-year sensitivity ranged from 62.3% to 76.0%, and specificity ranged from 56.0% to 87.5%. Because calibration was variable and long-term risk sets were small in some strata, these internally validated findings do not establish clinical utility.

## 4. DISCUSSION

In this large longitudinal memory-clinic cohort, plasma p-tau217 modified the prognostic association between chronological age and cognitive progression. Older age and higher plasma p-tau217 were each associated with faster worsening. However, the age-related risk gradient was smaller when p-tau217 was high. Results were consistent across time-to-event, absolute-risk, and longitudinal CDR-SB analyses. The prediction analyses also showed that the contribution of plasma p-tau217 differed across age groups.

Chronological age is a strong but non-specific predictor of cognitive decline. It captures variations in multiple biological and clinical processes, including comorbidity burden, cerebrovascular diseases, frailty, and mixed neuropathology.^33–36^ Recent work using brain-age, biological-age, and blood-biomarker approaches also show substantial heterogeneity in aging-related risk.^37,38^ Plasma p-tau217, by contrast, is a sensitive and specific blood biomarker for AD pathology and clinical progression, across symptomatic and preclinical disease stages.^8,39^ Our finding extend this work by showing plasma p-tau217 changes the prognostic implications of age.^40^ When p-tau217 was low, age retained substantial risk separation. When p-tau217 was high, progression risk was elevated in both age groups, leaving less additional separation by age. This is prognostic effect modification. It should not be interpreted as evidence that p-tau217 causally changes the effect of aging.

The Aβ-PET-anchored threshold analyses provide a clinically interpretable illustration of this interaction. The single plasma p-tau217 cutpoint of 0.546 pg/ml separated Aβ-positive from Aβ-negative individuals with high accuracy and was validated in the autopsy subset, although specificity was lower in the autopsy subset. Specificity was also less precise in the oldest PET age group. The dual-cutpoint approach further identified an intermediate group whose progression profile more closely resembled the high-risk group than the low-risk group, suggesting that clinically relevant risk may begin to rise before the highest biomarker threshold is reached. This finding is relevant for screening and trial enrichment, where binary classification may be insufficient and intermediate biomarker ranges may require closer follow-up or confirmatory testing.^41^

The prediction findings reinforce the need for age-contextualized interpretation. Adding p-tau217 produced the largest 5-year AUC gain below age 60 years. A low result provided less reassurance with advancing age, whereas the PPV of a high result tended to increase. This pattern is expected because predictive values depend on baseline risk and outcome prevalence.^42^ Calibration was variable in smaller strata. Therefore, the prediction analyses should be viewed as internally validated, supportive results rather than a clinical decision rule.

Clinically, in younger symptomatic or at-risk individuals, elevated plasma p-tau217 may identify a subgroup with clinically meaningful near-term progression risk despite otherwise favorable demographic profiles. Such individuals may be appropriate candidates for closer monitoring, confirmatory biomarker assessment, or enrichment in early-intervention trials. Conversely, in older individuals, low plasma p-tau217 may help avoid overestimation of AD-related progression risk although the reduced NPV in the oldest participants indicates that non-AD and mixed pathologies become increasingly important with advancing age. Previous studies suggest that older symptomatic individuals on the AD continuum may exhibit a lower tau burden at comparable level of clinical impairment, possibly because vascular, α-synuclein, TDP-43, and other age-related comorbid pathologies reduce the amount of AD-related tau pathology required to produce cognitive symptoms.^43,44^ Meanwhile, p-tau217 is among the earliest tau phosphorylation sites to become abnormal, but its trajectory may plateau as tau phosphorylation, cleavage, secretion, and aggregation may alter the relative expression of different tau species and increase the complementary relevance of markers such as p-tau205 and MTBR-tau243.^45–48^ Thus, plasma p-tau217 should not be interpreted in isolation, but in combination with age, clinical status, and the broader differential diagnosis.^49^

Strengths of this study include a large longitudinal clinical cohort with plasma p-tau217 measurements, clinically meaningful progression outcomes, and validation subsets with Aβ-PET imaging and autopsy data. Results were evaluated with complementary relative-risk, absolute-risk, incidence-rate, longitudinal, and prediction methods. Meanwhile, several limitations are important. The memory-clinic cohort was enriched for cognitive concerns and AD pathology, which may limit generalizability to population-based cohorts. Analyses were limited to participants who self-identified as Black or White. The analytic samples overlapped but were not identical, and follow-up was shorter in the high-p-tau217 groups. Death data were unavailable, so competing risks were not modeled. Vascular injury, frailty, kidney function, and non-AD neurodegenerative pathologies were not comprehensively modeled. Cutpoints may differ across assays and populations.^50^ Finally, some age-stratified prediction estimates, particularly at 10 years in the oldest participants, were based on limited long-term risk sets and should be interpreted as supportive.

In conclusion, our findings demonstrate that plasma p-tau217 modifies the prognostic association between chronological age and cognitive progression in this longitudinal memory-clinic cohort. Age strongly stratified risk when plasma p-tau217 was low. High p-tau217 identified elevated risk across age groups and attenuated the age-related gradients. These findings suggest that the prognostic significance of age varies by p-tau217 levels, supporting their integrated interpretation for prognosis, screening implementation, and trial enrichment.

## Data Availability

All data produced in the present study are available upon reasonable request to the authors

## ACKNOWLEDGMENTS/CONFLICTS/FUNDING SOURCES/CONSENT STATEMENT

## Acknowledgements

The authors are thankful to the Pitt-ADRC study participants, their family members and carers as well as study coordinators whose dedication and commitment were fundamental to the successful implementation of this study.

## Consent statement

All the studies were approved by the Institutional Review Board at the University of Pittsburgh. All subjects provided written informed consent.

## SUPPLEMENTARY FIGURE LEGENDS

**Figure S1.** Clinical diagnosis, age, and p-tau217 positivity across baseline severity. **A**, Baseline clinical diagnosis distribution by CDR-GS. Heat map showing the correspondence between the baseline CDR-GS and clinical diagnoses. Percentages indicate the proportion of participants with each CDR-GS category (0, 0.5, 1, 2, 3) classified as Control, MCI, AD dementia, or non-AD dementia. Cramér V summarizes association strength. **B**, Plasma p-tau217 levels by baseline clinical diagnosis. Boxes represent the interquartile ranges (IQRs), and center lines indicate the median. Group differences were assessed using the Kruskal-Wallis test followed by Dunn’s post hoc test with false discovery rate (FDR) adjustment. Fold change (fc) indicates the ratio of median values for pairwise comparisons. Asterisks indicate statistical significance (*p<0.05, **p<0.01, ***p<0.001, ****p<0.0001). **C**, Adjusted HR of p-tau217 for clinical diagnosis progression. Dose– response curve showing the adjusted HR for progression to a more severe clinical diagnosis according to plasma p-tau217 concentration. Risk increases with higher p-tau217 concentrations. **D**, Adjusted HR of age for CDR-GS progression. Dose–response curve showing the adjusted HR for CDR-GS progression according to age at blood draw. Risk increases with older age. **E-F**, Adjusted HR of p-tau217 for CDR-GS progression and clinical diagnosis progression by age group (<70 vs ≥70 years). The association between plasma p-tau217 and progression appears stronger in the younger cohort (age<70), as indicated by a steeper hazard curve. **G-H**, Prevalence of p-tau217 positivity (≥ 0.546 pg/mL) with binomial 95% CIs; counts shown as positive/total by age at 70 years (**G**) and 10-years age groups (**H**). Abbreviations: MCI, mild cognitive impairment; AD, Alzheimer disease dementia; CDR-GS, Clinical Dementia Rating Global Score; CI, confidence interval; HR, hazard ratio.

**Figure S2.** Performance of plasma p-tau217 for discriminating amyloid status using single and dual-cutpoint approaches. **A-D**, Plasma p-tau217 distribution by amyloid status using a single threshold. A, Plasma p-tau217 by amyloid status in the PET and autopsy cohorts. Boxes represent the interquartile range, and center lines indicate the median. The dashed green line marks the predicted cutpoint of 0.546 pg/mL. Samples classified as predicted negative are shown in blue, and those classified as predicted positive are shown in red. **B-C**, Confusion matrices for PET development and the displayed autopsy validation subset. Cells give counts and percentages of the displayed cohort. Actual status for amyloid-PET cohort is defined using Aβ-PET imaging obtained within 2-years of blood extraction (SUVR>1.346), and for autopsy cohort is defined using neuritic plaque burden. **D**, Sensitivity and specificity in broad and 10-year PET age groups with exact binomial 95% CIs. E-G, Plasma p-tau217 distribution by amyloid status using dual-threshold. **E**, Distributions with thresholds at 0.444 and 0.690 pg/mL. **F-G**, Three-category confusion matrices in the PET and displayed autopsy cohorts. Sensitivity and specificity printed for dual thresholds are calculated after excluding intermediate results. Abbreviations: A-, amyloid negative; A+, amyloid positive; Acc, accuracy; CI, confidence interval; Int, intermediate; IQR, interquartile range; Neg, negative; PET, positron emission tomography; Pos, positive; Sens, sensitivity; Spec, specificity.

**Figure S3.** Unadjusted progression risks and baseline clinical distributions across age-p-tau217 groups. **A**, Unadjusted Kaplan-Meier cumulative risks of CDR-GS progression at 2, 5, and 10 years. Bars show risks with 95% CIs; text gives older-minus-younger risk differences within p-tau217 groups, and the table gives numbers at risk. **B**, Baseline CDR-GS distribution across the 4 groups. **C**, Baseline diagnosis distribution across the 4 groups. Low and high p-tau217 were defined by 0.546 pg/mL; younger and older age by 70 years. Abbreviations: AD, Alzheimer disease dementia; CDR-GS, Clinical Dementia Rating global score; CI, confidence interval; MCI, mild cognitive impairment; pp, percentage points.

**Figure S4.** Age-stratified prediction performance of plasma p-tau217. **A**, Repeated-cross-validated time-dependent AUCs at 2, 5, and 10 years for the clinical and clinical-plus-p-tau217 Cox models. Error bars show bootstrap 95% CIs. **B**, Calibration of 5-year predicted risk against Kaplan-Meier observed risk across predicted-risk strata; the diagonal indicates perfect calibration. **C**, Age-stratified NPV and PPV at the fixed p-tau217 threshold of 0.546 pg/mL at 2, 5, and 10 years. Error bars show bootstrap 95% CIs. The clinical model included age, baseline CDR-GS, sex, education, APOE ε4, and self-reported race. Abbreviation: AUC, area under the curve; CDR-GS, Clinical Dementia Rating global score; CI, confidence interval; CV, cross-validation; KM, Kaplan-Meier; NPV, negative predictive value; PPV, positive predictive value.

