## Supplementary figures and images for "Association of age with clinical progression across plasma p-tau217 levels"

### Figure S1

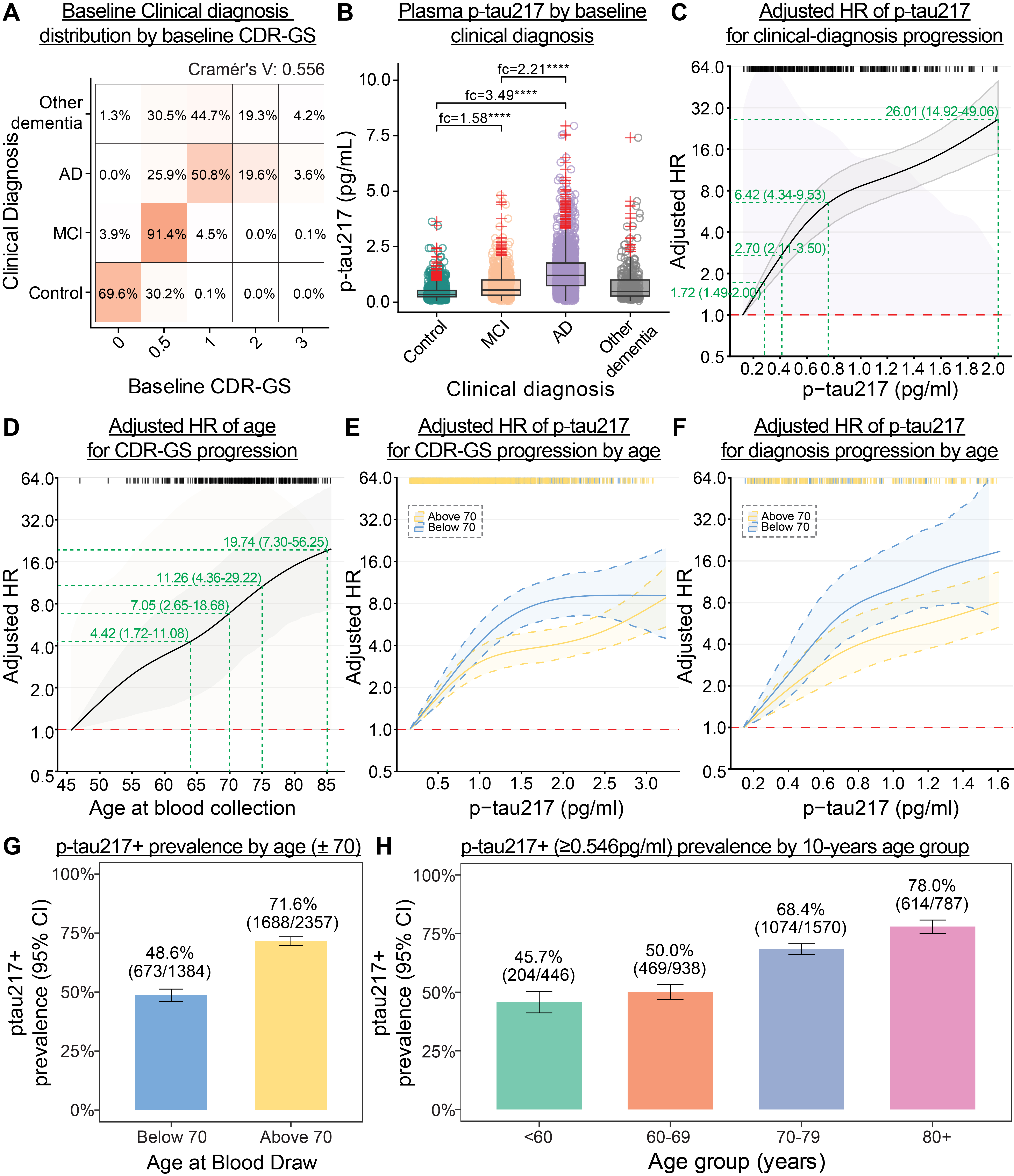

### Figure S2

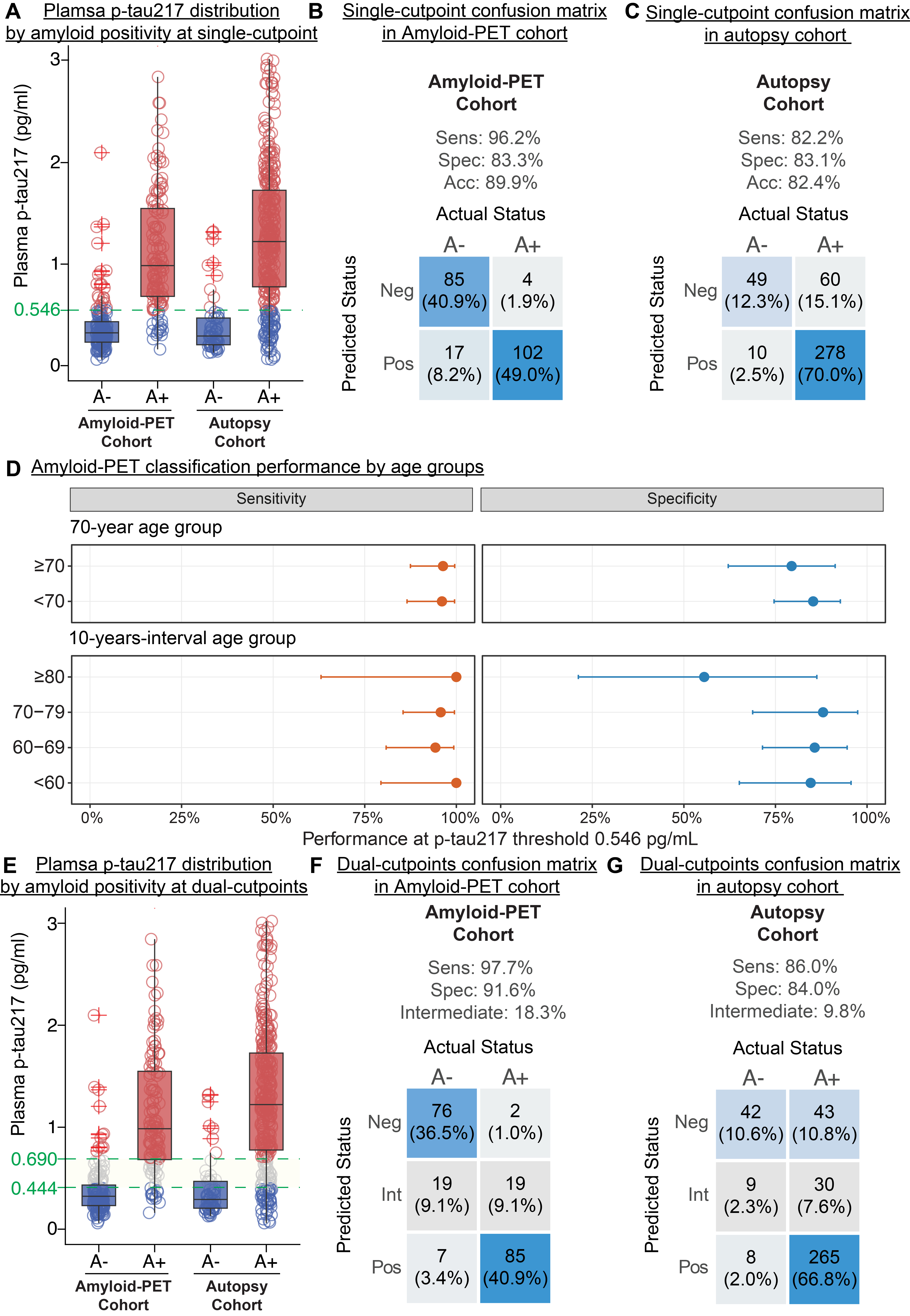

### Figure S3

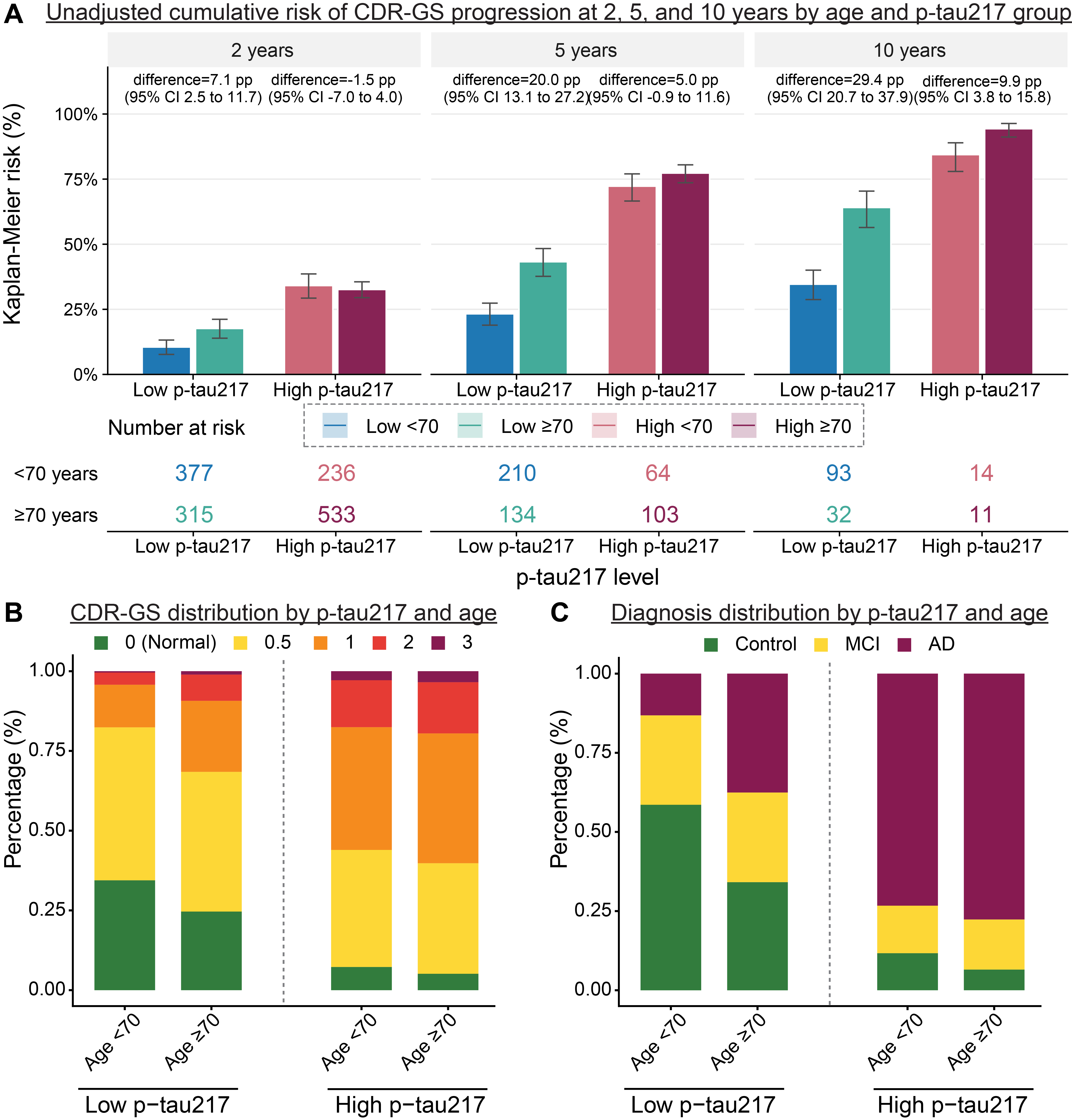

### Figure S4

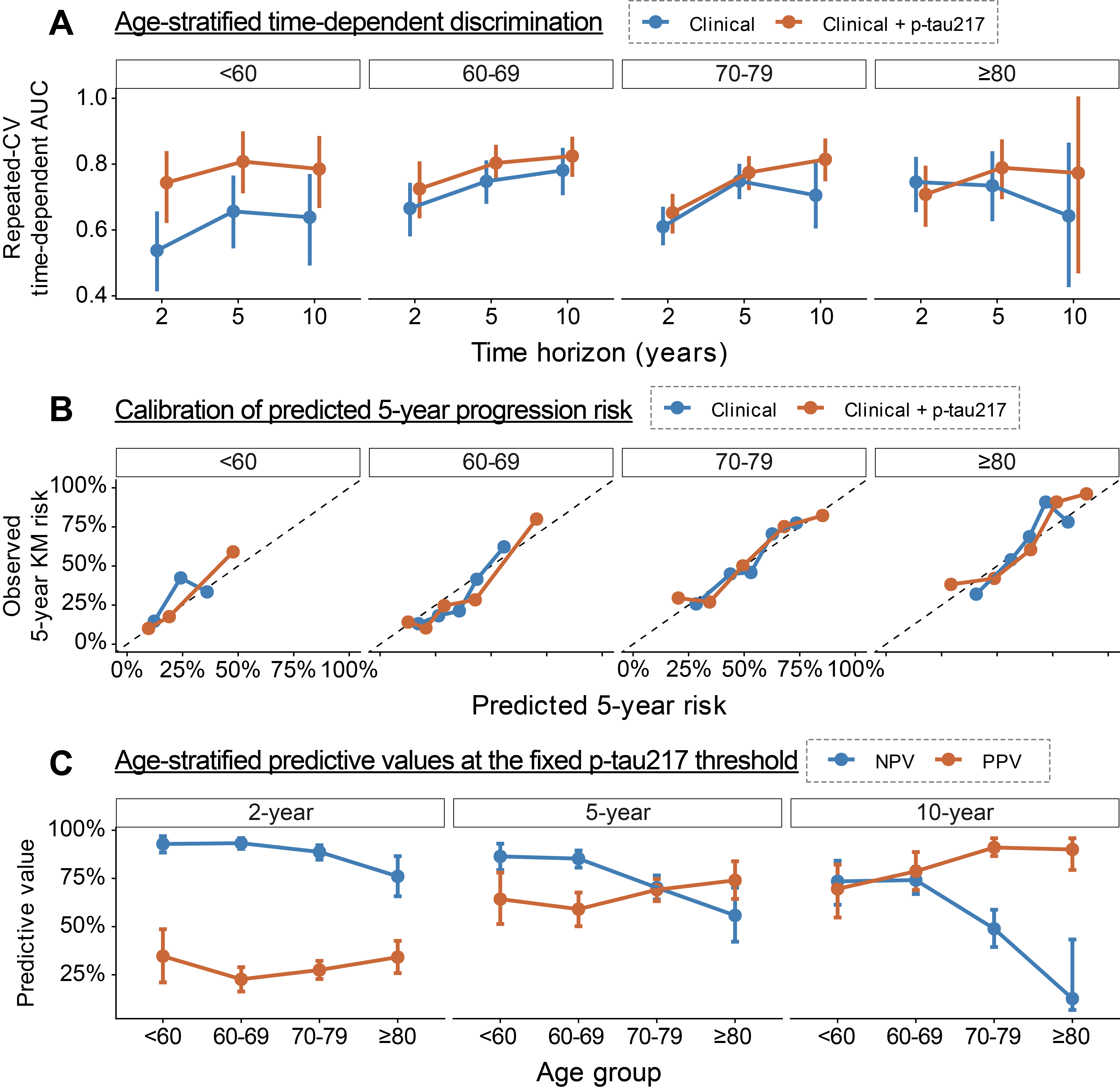
