## Supplementary material for "Association of age with clinical progression across plasma p-tau217 levels": Table S1

### Table S1. Baseline characteristics of the amyloid-status sub-cohorts

Table S1A. Non-autopsy Aβ-PET threshold-development cohort (Amyloid-PET sub-cohort)

| Variable | N | Overall | A- | A+ | p-value[1] |
|---|---|---|---|---|---|
| | | N = 208 | N = 102 | N = 106 | |
| **Sex** | 208 | | | | 0.97 |
| Female | | 92 (44%) | 45 (44%) | 47 (44%) | |
| Male | | 116 (56%) | 57 (56%) | 59 (56%) | |
| **Age** | 208 | 68.79 (62.19, 74.52) | 65.63 (59.80, 72.03) | 70.29 (64.05, 75.29) | <0.001 |
| **Race** | 208 | | | | >0.99 |
| B/AA | | 9 (4.3%) | 4 (3.9%) | 5 (4.7%) | |
| White | | 199 (96%) | 98 (96%) | 101 (95%) | |
| **Education** | 208 | | | | 0.056 |
| ≥13 yrs | | 173 (83%) | 90 (88%) | 83 (78%) | |
| < 13 yrs | | 35 (17%) | 12 (12%) | 23 (22%) | |
| **APOE ε4 carrier** | 208 | | | | <0.001 |
| Carrier | | 83 (40%) | 26 (25%) | 57 (54%) | |
| Non-carrier | | 125 (60%) | 76 (75%) | 49 (46%) | |
| **Clinical diagnosis** | 208 | | | | <0.001 |
| AD | | 58 (28%) | 8 (7.8%) | 50 (47%) | |
| Control | | 60 (29%) | 42 (41%) | 18 (17%) | |
| MCI | | 72 (35%) | 41 (40%) | 31 (29%) | |
| nonAD_Dem | | 18 (8.7%) | 11 (11%) | 7 (6.6%) | |
| **CDR-GS** | 208 | | | | <0.001 |
| 0 | | 37 (18%) | 23 (23%) | 14 (13%) | |
| 0.5 | | 129 (62%) | 68 (67%) | 61 (58%) | |
| 1 | | 35 (17%) | 7 (6.9%) | 28 (26%) | |
| 2 | | 7 (3.4%) | 4 (3.9%) | 3 (2.8%) | |
| **Incident AD** | 132 | 21 (16%) | 6 (7.2%) | 15 (31%) | <0.001 |
| **Diagnosis-progression** | 107 | 28 (26%) | 12 (18%) | 16 (41%) | 0.008 |
| **CDR-GS progression** | 163 | 50 (31%) | 14 (16%) | 36 (46%) | <0.001 |
| **p-tau217 (pg/mL)** | 208 | 0.63 (0.34, 1.07) | 0.34 (0.23, 0.48) | 1.02 (0.72, 1.62) | <0.001 |

Table S1B. Autopsy amyloid-status cohort (Autopsy sub-cohort)

| Variable | N | Overall | A- | A+ | p-value[1] |
|---|---|---|---|---|---|
| | | N = 397 | N = 59 | N = 338 | |
| **Sex** | 397 | | | | 0.024 |
| Female | | 202 (50.9%) | 22 (37.3%) | 180 (53.3%) | |
| Male | | 195 (49.1%) | 37 (62.7%) | 158 (46.7%) | |
| **Age** | 397 | 73.38 (64.72, 78.89) | 67.52 (60.06, 72.68) | 73.91 (65.39, 79.34) | <0.001 |
| **Race** | 397 | | | | 0.2 |
| B/AA | | 6 (1.5%) | 2 (3.4%) | 4 (1.2%) | |
| White | | 391 (98.5%) | 57 (96.6%) | 334 (98.8%) | |
| **Education** | 397 | | | | >0.9 |
| ≥13 yrs | | 257 (64.7%) | 38 (64.4%) | 219 (64.8%) | |
| < 13 yrs | | 140 (35.3%) | 21 (35.6%) | 119 (35.2%) | |
| **APOE ε4 carrier** | 397 | | | | <0.001 |
| Carrier | | 220 (55.4%) | 13 (22.0%) | 207 (61.2%) | |
| Non-carrier | | 177 (44.6%) | 46 (78.0%) | 131 (38.8%) | |
| **Clinical diagnosis** | 397 | | | | <0.001 |
| AD | | 271 (68.3%) | 11 (18.6%) | 260 (76.9%) | |
| Control | | 29 (7.3%) | 6 (10.2%) | 23 (6.8%) | |
| MCI | | 44 (11.1%) | 6 (10.2%) | 38 (11.2%) | |
| nonAD_Dem | | 53 (13.4%) | 36 (61.0%) | 17 (5.0%) | |
| **CDR-GS** | 397 | | | | 0.14 |
| 0 | | 26 (6.5%) | 8 (13.6%) | 18 (5.3%) | |
| 0.5 | | 121 (30.5%) | 15 (25.4%) | 106 (31.4%) | |
| 1 | | 179 (45.1%) | 27 (45.8%) | 152 (45.0%) | |
| 2 | | 60 (15.1%) | 9 (15.3%) | 51 (15.1%) | |
| 3 | | 11 (2.8%) | 0 (0.0%) | 11 (3.3%) | |
| **Incident AD** | 73 | 48 (65.8%) | 2 (16.7%) | 46 (75.4%) | <0.001 |
| **Diagnosis-progression** | 70 | 55 (78.6%) | 3 (30.0%) | 52 (86.7%) | <0.001 |
| **CDR-GS progression** | 353 | 270 (76.5%) | 36 (69.2%) | 234 (77.7%) | 0.2 |
| **p-tau217 (pg/mL)** | 397 | 1.14 (0.50, 1.63) | 0.29 (0.20, 0.47) | 1.22 (0.78, 1.73) | <0.001 |

*Continuous variables are reported as medians with interquartile ranges (IQRs), and categorical variables are summarized as counts and percentages. Aβ positivity was defined by an SUVR > 1.346 or via neuropathological assessment with a CERAD neuritic plaque score of moderate or frequent (NPNEUR 2 and 3).*

*[1]p-values were calculated with Wilcoxon rank-sum tests for continuous variables and Pearson's Chi-square or Fisher's exact tests for categorical variables, as appropriate.*

*Abbreviations: Aβ, amyloid-β; AD, Alzheimer disease dementia; B/AA, Black/African American; CDR-GS, Clinical Dementia Rating global score; MCI, mild cognitive impairment; PET, positron emission tomography*
