## Supplementary material for "Association of age with clinical progression across plasma p-tau217 levels": Table S2

**Table S2. Performance of the fixed p-tau217 threshold for Aβ-PET classification by age**

Table S2A. Sensitivity and specificity by age groups

| scheme | ge_group | metric | estimate | ci_low | ci_high | numerator | denominator | N |
|---|---|---|---|---|---|---|---|---|
| Binary age groups | <70 | Sensitivity | 0.961 | 0.865 | 0.995 | 49 | 51 | 119 |
| Binary age groups | <70 | Specificity | 0.853 | 0.746 | 0.927 | 58 | 68 | 119 |
| Binary age groups | >=70 | Sensitivity | 0.964 | 0.875 | 0.996 | 53 | 55 | 89 |
| Binary age groups | >=70 | Specificity | 0.794 | 0.621 | 0.913 | 27 | 34 | 89 |
| 10-year age groups | <60 | Sensitivity | 1.000 | 0.794 | 1.000 | 16 | 16 | 42 |
| 10-year age groups | <60 | Specificity | 0.846 | 0.651 | 0.956 | 22 | 26 | 42 |
| 10-year age groups | 60-69 | Sensitivity | 0.943 | 0.808 | 0.993 | 33 | 35 | 77 |
| 10-year age groups | 60-69 | Specificity | 0.857 | 0.715 | 0.946 | 36 | 42 | 77 |
| 10-year age groups | 70-79 | Sensitivity | 0.957 | 0.855 | 0.995 | 45 | 47 | 72 |
| 10-year age groups | 70-79 | Specificity | 0.880 | 0.688 | 0.975 | 22 | 25 | 72 |
| 10-year age groups | >=80 | Sensitivity | 1.000 | 0.631 | 1.000 | 8 | 8 | 17 |
| 10-year age groups | >=80 | Specificity | 0.556 | 0.212 | 0.863 | 5 | 9 | 17 |

Table S2B. Tests for performance heterogeneity across age groups

| scheme | metric | p_heterogeneity |
|---|---|---|
| Binary age groups | Sensitivity | 1.000 |
| Binary age groups | Specificity | 0.574 |
| 10-year age groups | Sensitivity | 1.000 |
| 10-year age groups | Specificity | 0.176 |

The fixed threshold was 0.546 pg/mL by maximizing the Youden-index and was not re-estimated by age. Table S2A reports exact binomial 95% CIs. Table S2B reports Fisher exact P values for heterogeneity across age groups.

Abbreviations: Aβ, amyloid-β; CI, confidence interval; PET, positron emission tomography.
