## Supplementary material for "Association of age with clinical progression across plasma p-tau217 levels": Table S3

### Table S3. Dual plasma p-tau217 thresholds and CDR-GS progression dual-threshold p-tau217 and age groups

Table S3A. Dual-threshold classification characteristics in the amyloid-PET cohort

| Lower threshold (pg/ml) | Upper threshold (pg/ml) | N | Sensitivity at lower threshold | Specificity at upper threshold | Intermediate proportion |
|---|---|---|---|---|---|
| 0.444 | 0.690 | 208 | 0.981 | 0.931 | 0.183 |

Table S3B. Adjusted CDR-GS progression by dual-threshold p-tau217 and age groups

| Comparison context | Joint group | N | Events | Adjusted HR | HR CI low | HR CI high | p value | Median years | Median CI low | Median CI high |
|---|---|---|---|---|---|---|---|---|---|---|
| Age comparison within low p-tau217 (Ref:<70) | Low p-tau217/Age<70 | 441 | 125 | - | - | - | - | 16.260 | 11.888 | 21.175 |
| | Low p-tau217/Age≥70 | 375 | 151 | 1.85 | 1.45 | 2.36 | 7.653E-07 | 7.513 | 6.018 | 9.366 |
| Age comparison within intermediate p-tau217 (Ref:<70) | Intermediate p-tau217/Age<70 | 132 | 48 | - | - | - | - | 8.381 | 7.565 | 13.498 |
| | Intermediate p-tau217/Age≥70 | 235 | 124 | 2.25 | 1.58 | 3.22 | 7.731E-06 | 3.992 | 3.869 | 4.367 |
| Age comparison within high p-tau217 (Ref:<70) | High p-tau217/Age<70 | 403 | 249 | - | - | - | - | 2.248 | 2.144 | 2.935 |
| | High p-tau217/Age≥70 | 928 | 567 | 1.00 | 0.86 | 1.16 | 0.956 | 2.913 | 2.456 | 3.012 |
| p-tau217 comparison within Below 70 (Ref: low) | Low p-tau217/Age<70 | 441 | 125 | - | - | - | - | 16.260 | 11.888 | 21.175 |
| | Intermediate p-tau217/Age<70 | 132 | 48 | 1.541 | 1.096 | 2.167 | 0.013 | 8.381 | 7.565 | 13.498 |

| Comparison context | Joint group | N | Events | Adjusted HR | HR CI low | HR CI high | p value | Median years | Median CI low | Median CI high |
|---|---|---|---|---|---|---|---|---|---|---|
| | High p-tau217/Age<70 | 403 | 249 | 4.704 | 3.691 | 5.996 | 6.638E-36 | 2.248 | 2.144 | 2.935 |
| p-tau217 comparison within Above 70 (Ref: low) | Low p-tau217/Age≥70 | 375 | 151 | - | - | - | - | 7.513 | 6.018 | 9.366 |
| | Intermediate p-tau217/Age≥70 | 235 | 124 | 1.863 | 1.461 | 2.376 | 5.263E-07 | 3.992 | 3.869 | 4.367 |
| | High p-tau217/Age≥70 | 928 | 567 | 3.059 | 2.519 | 3.714 | 1.531E-29 | 2.913 | 2.456 | 3.012 |

Table S3A reports sensitivity at the lower threshold (0.444 pg/mL), specificity at the upper threshold (0.690 pg/mL), and the intermediate proportion in all 208 PET participants. These differ from the apparent sensitivity and specificity calculated after excluding intermediate results in Supplementary Figure 2F. Table S3B reports Cox HRs adjusted for sex, education, APOE ε4, and self-reported race, with Kaplan-Meier median progression-free times. Dashes indicate reference groups.

Abbreviations: CDR-GS, Clinical Dementia Rating global score; CI, confidence interval; HR, hazard ratio; PET, positron emission tomography.
