## Supplementary material for "Association of age with clinical progression across plasma p-tau217 levels": Table S4

### Table S4. CDR-GS progression by the single p-tau217 threshold and age

Table S4A. Incidence rates and incidence rate ratios

| Group | N | Events | IR per 100 PY (95%CI) | IRR (95% CI) |
|---|---|---|---|---|
| Low p-tau217/Age<70 | 509 | 148 | 5.04 (4.26-5.92) | 1.00 (1.00-1.00) |
| Low p-tau217/Age≥70 | 471 | 201 | 10.43 (9.04-11.97) | 2.07 (1.68-2.56) |
| High p-tau217/Age<70 | 467 | 274 | 21.15 (18.72-23.81) | 4.20 (3.44-5.14) |
| High p-tau217/Age≥70 | 1067 | 641 | 24.14 (22.31-26.08) | 4.79 (4.02-5.75) |

Table S4B. Adjusted hazard ratios (HR) and median progression-free times

| Comparison group | Reference group | N | Events | HR | HR | | P value | Comparison median | | | Reference median | | |
|---|---|---|---|---|---|---|---|---|---|---|---|---|---|
| | | | | | CI Low | CI High | | Years | CI Low | CI High | Years | CI Low | CI High |
| Low p-tau217/Above 70 years | Low p-tau217/ Below 70 years | 980 | 349 | 1.97 | 1.58 | 2.45 | 1.582E-09 | 6.223 | 5.782 | 8.630 | 14.897 | 11.888 | 19.006 |
| High p-tau217/Above 70 years | High p-tau217/ Below 70 years | 1,534 | 915 | 1.10 | 0.95 | 1.26 | 0.216 | 2.979 | 2.836 | 3.047 | 2.880 | 2.248 | 3.014 |
| High p-tau217/Below 70 years | Low p-tau217/ Below 70 years | 976 | 422 | 3.88 | 3.09 | 4.86 | 5.629E-32 | 2.880 | 2.248 | 3.014 | 14.897 | 11.888 | 19.006 |
| High p-tau217/Above 70 years | Low p-tau217/ Above 70 years | 1,538 | 842 | 2.51 | 2.12 | 2.97 | 2.171E-26 | 2.979 | 2.836 | 3.047 | 6.223 | 5.782 | 8.630 |

Low and high p-tau217 were defined as < 0.546 and ≥ 0.546 pg/mL. Younger and older age were defined as < 70 and ≥ 70 years. Table S4A reports incidence per 100 person-years and incidence rate ratios. Table S4B reports Cox HRs adjusted for sex, education, APOE ε4, and self-reported race, with Kaplan-Meier median progression-free times.
Abbreviations: CDR-GS, Clinical Dementia Rating global score; CI, confidence interval; HR, hazard ratio; IRR, incidence rate ratio; PY, person-years.
