## Supplementary material for "Association of age with clinical progression across plasma p-tau217 levels": Table S5

### Table S5. Clinical diagnosis progression by the single p-tau217 threshold and age

Table S5A. Incidence rates and incidence rate ratios

| Group | N | Events | IR per 100 PY (95%CI) | IRR (95% CI) |
|---|---|---|---|---|
| Low p-tau217/Age<70 | 388 | 62 | 2.37 (1.82-3.04) | 1.00 (1.00-1.00) |
| Low p-tau217/Age≥70 | 296 | 89 | 6.06 (4.86-7.45) | 2.55(1.85-3.55) |
| High p-tau217/Age<70 | 129 | 62 | 10.66 (8.17-13.66) | 4.49 (3.16-6.40) |
| High p-tau217/Age≥70 | 269 | 164 | 18.05 (15.40-21.04) | 7.61(5.72-10.37) |

Table S5B. Adjusted hazard ratios and median progression-free times

| Comparison group | Reference group | N | Events | HR | | | P value | Comparison median | | | Reference median | | |
|---|---|---|---|---|---|---|---|---|---|---|---|---|---|
| | | | | | CI Low | CI High | | Years | CI Low | CI High | Years | CI Low | CI High |
| Low p-tau217/Above 70 years | Low p-tau217/ Below 70 years | 684 | 151 | 2.93 | 2.08 | 4.12 | 6.285E-10 | 13.153 | 9.240 | 16.991 | 21.054 | 19.001 | |
| High p-tau217/Above 70 years | High p-tau217/ Below 70 years | 398 | 226 | 1.93 | 1.43 | 2.61 | 1.677E-05 | 3.838 | 3.146 | 4.068 | 6.007 | 4.775 | 7.989 |
| High p-tau217/Below 70 years | Low p-tau217/ Below 70 years | 517 | 124 | 5.18 | 3.47 | 7.73 | 9.197E-16 | 6.007 | 4.775 | 7.989 | 21.054 | 19.001 | |
| High p-tau217/Above 70 years | Low p-tau217/ Above 70 years | 565 | 253 | 3.37 | 2.54 | 4.46 | 2.702E-17 | 3.838 | 3.146 | 4.068 | 13.153 | 9.240 | 16.991 |

*Clinical diagnosis progression was CU to MCI or AD dementia, or MCI to AD dementia. Low and high p-tau217 were defined by 0.546 pg/mL; younger and older age by 70 years. Table S5A reports incidence per 100 person-years and incidence rate ratios. Table S5B reports Cox HRs adjusted for sex, education, APOE ε4, and self-reported race, with Kaplan-Meier median progression-free times.*

*Abbreviations: AD, Alzheimer disease; CI, confidence interval; CU, cognitively unimpaired; HR, hazard ratio; IRR, incidence rate ratio; MCI, mild cognitive impairment; PY, person-years.*
