## Supplementary material for "Association of age with clinical progression across plasma p-tau217 levels": Table S6

**Table S6. Covariate-standardized 5-year risk of CDR-GS progression**

Table S6A. Standardized 5-year risks by p-tau217 and age group

| Group | Risk | CI low | CI high | boot_n | risk_pct | ci_low_pct | ci_high_pct |
|---|---|---|---|---|---|---|---|
| Low p-tau217/Age<70 | 0.257 | 0.218 | 0.297 | 1000 | 25.670 | 21.825 | 29.660 |
| Low p-tau217/Age≥70 | 0.450 | 0.406 | 0.495 | 1000 | 44.968 | 40.582 | 49.481 |
| High p-tau217/Age<70 | 0.715 | 0.665 | 0.760 | 1000 | 71.474 | 66.472 | 76.040 |
| High p-tau217/Age≥70 | 0.748 | 0.717 | 0.775 | 1000 | 74.778 | 71.723 | 77.500 |

Table S6B. Older-minus-younger standardized risk differences

| contrast | estimate | ci_low | ci_high |
|---|---|---|---|
| Older - younger within low p-tau217 | 19.299 | 13.542 | 24.984 |
| Older - younger within high p-tau217 | 3.304 | -1.386 | 8.328 |

Table S6C. Analytic sample and follow-up by group

| Group | N | events | median_followup | q1_followup | q3_followup | median_visits |
|---|---|---|---|---|---|---|
| Low p-tau217/Age<70 | 509 | 148 | 4.063 | 1.982 | 7.992 | 4 |
| Low p-tau217/Age≥70 | 471 | 201 | 2.984 | 1.399 | 5.804 | 4 |
| High p-tau217/Age<70 | 467 | 274 | 2.004 | 1.113 | 3.155 | 3 |
| High p-tau217/Age≥70 | 1067 | 641 | 1.999 | 1.080 | 3.091 | 3 |

Risks were standardized over the covariate distribution of the CDR-GS progression cohort from a Cox model adjusted for sex, education, APOE ε4, and self-reported race. CIs used 1,000 bootstrap resamples. Table S6C describes differential follow-up across groups.

Abbreviations: CDR-GS, Clinical Dementia Rating global score; CI, confidence interval
