## Supplementary material for "Association of age with clinical progression across plasma p-tau217 levels": Table S7

**Table S7. Five-year piecewise-linear CDR-SB sensitivity analysis**

Table S7A. Comparison of linear and piecewise-linear time models

| model | AIC | BIC | logLik | comparison_p |
|---|---|---|---|---|
| Linear time, 0-5 years | 38509.733 | 38587.482 | -19243.866 | |
| Piecewise time, knot at 2 years | 38396.507 | 38502.528 | -19183.254 | 2.92E-25 |

Table S7B. Estimated CDR-SB slopes by time segment and group

| group | age | segment | slope | ci_low | ci_high |
|---|---|---|---|---|---|
| High p-tau217/Age<70 | <70 | 0-2 years | 1.185 | 1.055 | 1.314 |
| High p-tau217/Age<70 | <70 | >2-5 years | 1.700 | 1.535 | 1.865 |
| High p-tau217/Age≥70 | >=70 | 0-2 years | 1.137 | 1.051 | 1.224 |
| High p-tau217/Age≥70 | >=70 | >2-5 years | 1.633 | 1.525 | 1.740 |
| Low p-tau217/Age<70 | <70 | 0-2 years | 0.270 | 0.147 | 0.392 |
| Low p-tau217/Age<70 | <70 | >2-5 years | 0.286 | 0.145 | 0.426 |
| Low p-tau217/Age≥70 | >=70 | 0-2 years | 0.481 | 0.352 | 0.610 |
| Low p-tau217/Age≥70 | >=70 | >2-5 years | 0.564 | 0.420 | 0.708 |

Table S7C. Time-by-age-by-p-tau217 interaction terms

| term | Estimate | Std. Error | df | t value | Pr(>\|t\|) |
|---|---|---|---|---|---|
| time_0_2:ptau_fhigh:age_f>=70 | -0.259 | 0.121 | 2457.019 | -2.144 | 0.032 |
| time_after2:ptau_fhigh:age_f>=70 | -0.346 | 0.144 | 3605.431 | -2.409 | 0.016 |

Models used maximum likelihood and included fixed effects for p-tau217 group, age group, time segment, and all interactions. A participant-specific random intercept and slope were used when supported; a random-intercept model was used after a singular fit. The piecewise model had a knot at 2 years. Slopes are CDR-SB points per year.
Abbreviations: CDR-SB, Clinical Dementia Rating Sum of Boxes; CI, confidence interval.
