## Supplementary material for "Association of age with clinical progression across plasma p-tau217 levels": Table S8

**Table S8. Age-stratified prediction performance for CDR-GS progression**

Table S8A. Repeated-cross-validated time-dependent discrimination

| Age group | Time (years) | N | events | AUC (base) | base_low | base_high | AUC (aug) | aug_low | aug_high | delta | delta_low | delta_high | delta_boot_sd | boot_n |
|---|---|---|---|---|---|---|---|---|---|---|---|---|---|---|
| <60 | 2 | 184 | 64 | 0.538 | 0.419 | 0.651 | 0.744 | 0.627 | 0.834 | 0.206 | 0.093 | 0.312 | 0.057 | 1000 |
| <60 | 5 | 184 | 64 | 0.657 | 0.550 | 0.760 | 0.808 | 0.717 | 0.894 | 0.151 | 0.073 | 0.238 | 0.042 | 1000 |
| <60 | 10 | 184 | 64 | 0.639 | 0.498 | 0.764 | 0.785 | 0.673 | 0.880 | 0.147 | 0.071 | 0.229 | 0.040 | 1000 |
| 60-69 | 2 | 477 | 166 | 0.666 | 0.586 | 0.738 | 0.725 | 0.641 | 0.803 | 0.059 | -0.004 | 0.121 | 0.031 | 1000 |
| 60-69 | 5 | 477 | 166 | 0.748 | 0.685 | 0.806 | 0.803 | 0.752 | 0.853 | 0.056 | 0.015 | 0.098 | 0.021 | 1000 |
| 60-69 | 10 | 477 | 166 | 0.781 | 0.711 | 0.844 | 0.825 | 0.767 | 0.878 | 0.043 | -0.003 | 0.094 | 0.025 | 1000 |
| 70-79 | 2 | 630 | 320 | 0.610 | 0.559 | 0.665 | 0.652 | 0.595 | 0.704 | 0.042 | -0.004 | 0.089 | 0.024 | 1000 |
| 70-79 | 5 | 630 | 320 | 0.749 | 0.699 | 0.795 | 0.774 | 0.727 | 0.818 | 0.026 | -0.017 | 0.072 | 0.022 | 1000 |
| 70-79 | 10 | 630 | 320 | 0.705 | 0.610 | 0.799 | 0.814 | 0.754 | 0.872 | 0.109 | 0.044 | 0.178 | 0.035 | 1000 |
| >=80 | 2 | 210 | 115 | 0.746 | 0.660 | 0.816 | 0.708 | 0.615 | 0.790 | -0.037 | -0.115 | 0.041 | 0.040 | 1000 |
| >=80 | 5 | 210 | 115 | 0.735 | 0.632 | 0.833 | 0.789 | 0.699 | 0.870 | 0.054 | -0.037 | 0.145 | 0.046 | 1000 |
| >=80 | 10 | 210 | 115 | 0.643 | 0.432 | 0.860 | 0.773 | 0.474 | 1.000 | 0.130 | -0.047 | 0.285 | 0.085 | 858 |

Table S8B. Prediction error by Brier score

| age_group | time_yr | N | brier_base | brier_aug | delta_brier |
|---|---|---|---|---|---|
| <60 | 2 | 184 | 0.136 | 0.120 | -0.015 |
| <60 | 5 | 184 | 0.206 | 0.151 | -0.055 |
| <60 | 10 | 184 | 0.238 | 0.186 | -0.052 |
| 60-69 | 2 | 477 | 0.106 | 0.096 | -0.010 |
| 60-69 | 5 | 477 | 0.181 | 0.151 | -0.030 |
| 60-69 | 10 | 477 | 0.197 | 0.170 | -0.027 |
| 70-79 | 2 | 630 | 0.161 | 0.158 | -0.003 |
| 70-79 | 5 | 630 | 0.204 | 0.192 | -0.012 |
| 70-79 | 10 | 630 | 0.173 | 0.149 | -0.024 |
| >=80 | 2 | 210 | 0.189 | 0.185 | -0.004 |
| >=80 | 5 | 210 | 0.195 | 0.181 | -0.014 |
| >=80 | 10 | 210 | 0.118 | 0.107 | -0.011 |

Table S8C. Calibration data by predicted-risk stratum

| bin | N | predicted | observed | model | time_yr | age_group |
|---|---|---|---|---|---|---|
| 1 | 62 | 0.042 | 0.101 | Clinical | 2 | <60 |
| 2 | 61 | 0.086 | 0.226 | Clinical | 2 | <60 |
| 3 | 61 | 0.135 | 0.138 | Clinical | 2 | <60 |
| 1 | 62 | 0.030 | 0.049 | Clinical + p-tau217 | 2 | <60 |
| 2 | 61 | 0.061 | 0.123 | Clinical + p-tau217 | 2 | <60 |
| 3 | 61 | 0.184 | 0.289 | Clinical + p-tau217 | 2 | <60 |
| 1 | 62 | 0.122 | 0.147 | Clinical | 5 | <60 |
| 2 | 61 | 0.241 | 0.424 | Clinical | 5 | <60 |
| 3 | 61 | 0.360 | 0.334 | Clinical | 5 | <60 |
| 1 | 62 | 0.097 | 0.101 | Clinical + p-tau217 | 5 | <60 |
| 2 | 61 | 0.190 | 0.176 | Clinical + p-tau217 | 5 | <60 |
| 3 | 61 | 0.477 | 0.591 | Clinical + p-tau217 | 5 | <60 |
| 1 | 62 | 0.215 | 0.332 | Clinical | 10 | <60 |
| 2 | 61 | 0.403 | 0.460 | Clinical | 10 | <60 |
| 3 | 61 | 0.564 | 0.441 | Clinical | 10 | <60 |
| 1 | 62 | 0.190 | 0.242 | Clinical + p-tau217 | 10 | <60 |
| 2 | 61 | 0.356 | 0.228 | Clinical + p-tau217 | 10 | <60 |
| 3 | 61 | 0.716 | 0.698 | Clinical + p-tau217 | 10 | <60 |
| 1 | 96 | 0.059 | 0.043 | Clinical | 2 | 60-69 |
| 2 | 96 | 0.094 | 0.099 | Clinical | 2 | 60-69 |
| 3 | 95 | 0.133 | 0.122 | Clinical | 2 | 60-69 |
| 4 | 95 | 0.169 | 0.125 | Clinical | 2 | 60-69 |
| 5 | 95 | 0.233 | 0.248 | Clinical | 2 | 60-69 |
| 1 | 96 | 0.039 | 0.054 | Clinical + p-tau217 | 2 | 60-69 |
| 2 | 96 | 0.066 | 0.076 | Clinical + p-tau217 | 2 | 60-69 |
| 3 | 95 | 0.096 | 0.092 | Clinical + p-tau217 | 2 | 60-69 |
| 4 | 95 | 0.154 | 0.047 | Clinical + p-tau217 | 2 | 60-69 |
| 5 | 95 | 0.315 | 0.364 | Clinical + p-tau217 | 2 | 60-69 |

| bin | N | predicted | observed | model | time_yr | age_group |
|---|---|---|---|---|---|---|
| 1 | 96 | 0.173 | 0.131 | Clinical | 5 | 60-69 |
| 2 | 96 | 0.263 | 0.182 | Clinical | 5 | 60-69 |
| 3 | 95 | 0.357 | 0.213 | Clinical | 5 | 60-69 |
| 4 | 95 | 0.436 | 0.415 | Clinical | 5 | 60-69 |
| 5 | 95 | 0.558 | 0.622 | Clinical | 5 | 60-69 |
| 1 | 96 | 0.126 | 0.141 | Clinical + p-tau217 | 5 | 60-69 |
| 2 | 96 | 0.206 | 0.104 | Clinical + p-tau217 | 5 | 60-69 |
| 3 | 95 | 0.289 | 0.247 | Clinical + p-tau217 | 5 | 60-69 |
| 4 | 95 | 0.428 | 0.284 | Clinical + p-tau217 | 5 | 60-69 |
| 5 | 95 | 0.704 | 0.800 | Clinical + p-tau217 | 5 | 60-69 |
| 1 | 96 | 0.298 | 0.167 | Clinical | 10 | 60-69 |
| 2 | 96 | 0.435 | 0.394 | Clinical | 10 | 60-69 |
| 3 | 95 | 0.562 | 0.361 | Clinical | 10 | 60-69 |
| 4 | 95 | 0.657 | 0.605 | Clinical | 10 | 60-69 |
| 5 | 95 | 0.781 | 0.829 | Clinical | 10 | 60-69 |
| 1 | 96 | 0.246 | 0.243 | Clinical + p-tau217 | 10 | 60-69 |
| 2 | 96 | 0.383 | 0.185 | Clinical + p-tau217 | 10 | 60-69 |
| 3 | 95 | 0.509 | 0.411 | Clinical + p-tau217 | 10 | 60-69 |
| 4 | 95 | 0.686 | 0.494 | Clinical + p-tau217 | 10 | 60-69 |
| 5 | 95 | 0.911 | 0.969 | Clinical + p-tau217 | 10 | 60-69 |
| 1 | 126 | 0.103 | 0.117 | Clinical | 2 | 70-79 |
| 2 | 126 | 0.170 | 0.211 | Clinical | 2 | 70-79 |
| 3 | 126 | 0.217 | 0.171 | Clinical | 2 | 70-79 |
| 4 | 126 | 0.273 | 0.230 | Clinical | 2 | 70-79 |
| 5 | 126 | 0.351 | 0.299 | Clinical | 2 | 70-79 |
| 1 | 126 | 0.066 | 0.168 | Clinical + p-tau217 | 2 | 70-79 |
| 2 | 126 | 0.118 | 0.062 | Clinical + p-tau217 | 2 | 70-79 |
| 3 | 126 | 0.184 | 0.155 | Clinical + p-tau217 | 2 | 70-79 |
| 4 | 126 | 0.290 | 0.292 | Clinical + p-tau217 | 2 | 70-79 |

| bin | N | predicted | observed | model | time_yr | age_group |
|---|---|---|---|---|---|---|
| 5 | 126 | 0.449 | 0.344 | Clinical + p-tau217 | 2 | 70-79 |
| 1 | 126 | 0.284 | 0.258 | Clinical | 5 | 70-79 |
| 2 | 126 | 0.438 | 0.449 | Clinical | 5 | 70-79 |
| 3 | 126 | 0.530 | 0.459 | Clinical | 5 | 70-79 |
| 4 | 126 | 0.627 | 0.705 | Clinical | 5 | 70-79 |
| 5 | 126 | 0.735 | 0.775 | Clinical | 5 | 70-79 |
| 1 | 126 | 0.204 | 0.295 | Clinical + p-tau217 | 5 | 70-79 |
| 2 | 126 | 0.344 | 0.270 | Clinical + p-tau217 | 5 | 70-79 |
| 3 | 126 | 0.494 | 0.501 | Clinical + p-tau217 | 5 | 70-79 |
| 4 | 126 | 0.681 | 0.752 | Clinical + p-tau217 | 5 | 70-79 |
| 5 | 126 | 0.853 | 0.823 | Clinical + p-tau217 | 5 | 70-79 |
| 1 | 126 | 0.462 | 0.547 | Clinical | 10 | 70-79 |
| 2 | 126 | 0.659 | 0.730 | Clinical | 10 | 70-79 |
| 3 | 126 | 0.757 | 0.749 | Clinical | 10 | 70-79 |
| 4 | 126 | 0.842 | 0.839 | Clinical | 10 | 70-79 |
| 5 | 126 | 0.915 | 0.887 | Clinical | 10 | 70-79 |
| 1 | 126 | 0.377 | 0.470 | Clinical + p-tau217 | 10 | 70-79 |
| 2 | 126 | 0.585 | 0.586 | Clinical + p-tau217 | 10 | 70-79 |
| 3 | 126 | 0.757 | 0.778 | Clinical + p-tau217 | 10 | 70-79 |
| 4 | 126 | 0.906 | 0.941 | Clinical + p-tau217 | 10 | 70-79 |
| 5 | 126 | 0.978 |  | Clinical + p-tau217 | 10 | 70-79 |
| 1 | 42 | 0.156 | 0.101 | Clinical | 2 | >=80 |
| 2 | 42 | 0.235 | 0.134 | Clinical | 2 | >=80 |
| 3 | 42 | 0.285 | 0.292 | Clinical | 2 | >=80 |
| 4 | 42 | 0.337 | 0.471 | Clinical | 2 | >=80 |
| 5 | 42 | 0.428 | 0.535 | Clinical | 2 | >=80 |
| 1 | 42 | 0.098 | 0.246 | Clinical + p-tau217 | 2 | >=80 |
| 2 | 42 | 0.181 | 0.130 | Clinical + p-tau217 | 2 | >=80 |
| 3 | 42 | 0.269 | 0.101 | Clinical + p-tau217 | 2 | >=80 |

| bin | N | predicted | observed | model | time_yr | age_group |
|---|---|---|---|---|---|---|
| 4 | 42 | 0.354 | 0.457 | Clinical + p-tau217 | 2 | >=80 |
| 5 | 42 | 0.517 | 0.599 | Clinical + p-tau217 | 2 | >=80 |
| 1 | 42 | 0.406 | 0.320 | Clinical | 5 | >=80 |
| 2 | 42 | 0.562 | 0.540 | Clinical | 5 | >=80 |
| 3 | 42 | 0.645 | 0.687 | Clinical | 5 | >=80 |
| 4 | 42 | 0.719 | 0.909 | Clinical | 5 | >=80 |
| 5 | 42 | 0.819 | 0.782 | Clinical | 5 | >=80 |
| 1 | 42 | 0.292 | 0.382 | Clinical + p-tau217 | 5 | >=80 |
| 2 | 42 | 0.487 | 0.421 | Clinical + p-tau217 | 5 | >=80 |
| 3 | 42 | 0.651 | 0.604 | Clinical + p-tau217 | 5 | >=80 |
| 4 | 42 | 0.767 | 0.909 | Clinical + p-tau217 | 5 | >=80 |
| 5 | 42 | 0.902 | 0.961 | Clinical + p-tau217 | 5 | >=80 |
| 1 | 42 | 0.618 | 0.869 | Clinical | 10 | >=80 |
| 2 | 42 | 0.787 |  | Clinical | 10 | >=80 |
| 3 | 42 | 0.856 | 0.766 | Clinical | 10 | >=80 |
| 4 | 42 | 0.907 |  | Clinical | 10 | >=80 |
| 5 | 42 | 0.957 |  | Clinical | 10 | >=80 |
| 1 | 42 | 0.509 | 0.811 | Clinical + p-tau217 | 10 | >=80 |
| 2 | 42 | 0.750 |  | Clinical + p-tau217 | 10 | >=80 |
| 3 | 42 | 0.888 | 0.871 | Clinical + p-tau217 | 10 | >=80 |
| 4 | 42 | 0.951 |  | Clinical + p-tau217 | 10 | >=80 |
| 5 | 42 | 0.990 |  | Clinical + p-tau217 | 10 | >=80 |

Table S8D. Prognostic performance of the fixed p-tau217 threshold

| age_group | time_yr | metric | estimate | ci_low | ci_high |
|---|---|---|---|---|---|
| <60 | 2 | NPV | 0.929 | 0.884 | 0.969 |
| <60 | 2 | PPV | 0.347 | 0.210 | 0.486 |
| <60 | 2 | Sensitivity | 0.659 | 0.475 | 0.829 |
| <60 | 2 | Specificity | 0.776 | 0.706 | 0.841 |
| <60 | 5 | NPV | 0.864 | 0.794 | 0.930 |

| age_group | time_yr | metric | estimate | ci_low | ci_high |
|---|---|---|---|---|---|
| <60 | 5 | PPV | 0.643 | 0.513 | 0.780 |
| <60 | 5 | Sensitivity | 0.623 | 0.492 | 0.756 |
| <60 | 5 | Specificity | 0.875 | 0.806 | 0.948 |
| <60 | 10 | NPV | 0.735 | 0.613 | 0.842 |
| <60 | 10 | PPV | 0.696 | 0.547 | 0.822 |
| <60 | 10 | Sensitivity | 0.495 | 0.373 | 0.624 |
| <60 | 10 | Specificity | 0.884 | 0.792 | 0.968 |
| 60-69 | 2 | NPV | 0.933 | 0.903 | 0.959 |
| 60-69 | 2 | PPV | 0.227 | 0.163 | 0.290 |
| 60-69 | 2 | Sensitivity | 0.671 | 0.537 | 0.786 |
| 60-69 | 2 | Specificity | 0.671 | 0.626 | 0.716 |
| 60-69 | 5 | NPV | 0.853 | 0.806 | 0.895 |
| 60-69 | 5 | PPV | 0.590 | 0.502 | 0.677 |
| 60-69 | 5 | Sensitivity | 0.719 | 0.627 | 0.801 |
| 60-69 | 5 | Specificity | 0.771 | 0.724 | 0.816 |
| 60-69 | 10 | NPV | 0.741 | 0.669 | 0.810 |
| 60-69 | 10 | PPV | 0.787 | 0.689 | 0.887 |
| 60-69 | 10 | Sensitivity | 0.662 | 0.580 | 0.765 |
| 60-69 | 10 | Specificity | 0.834 | 0.773 | 0.902 |
| 70-79 | 2 | NPV | 0.888 | 0.847 | 0.922 |
| 70-79 | 2 | PPV | 0.275 | 0.228 | 0.322 |
| 70-79 | 2 | Sensitivity | 0.764 | 0.683 | 0.833 |
| 70-79 | 2 | Specificity | 0.480 | 0.435 | 0.525 |
| 70-79 | 5 | NPV | 0.702 | 0.640 | 0.765 |
| 70-79 | 5 | PPV | 0.691 | 0.632 | 0.747 |
| 70-79 | 5 | Sensitivity | 0.760 | 0.706 | 0.815 |
| 70-79 | 5 | Specificity | 0.627 | 0.563 | 0.687 |
| 70-79 | 10 | NPV | 0.489 | 0.394 | 0.587 |
| 70-79 | 10 | PPV | 0.911 | 0.866 | 0.959 |

| age_group | time_yr | metric | estimate | ci_low | ci_high |
|---|---|---|---|---|---|
| 70-79 | 10 | Sensitivity | 0.705 | 0.648 | 0.762 |
| 70-79 | 10 | Specificity | 0.798 | 0.706 | 0.916 |
| >=80 | 2 | NPV | 0.761 | 0.657 | 0.866 |
| >=80 | 2 | PPV | 0.342 | 0.258 | 0.426 |
| >=80 | 2 | Sensitivity | 0.718 | 0.601 | 0.836 |
| >=80 | 2 | Specificity | 0.395 | 0.313 | 0.480 |
| >=80 | 5 | NPV | 0.558 | 0.422 | 0.702 |
| >=80 | 5 | PPV | 0.740 | 0.644 | 0.839 |
| >=80 | 5 | Sensitivity | 0.739 | 0.663 | 0.820 |
| >=80 | 5 | Specificity | 0.560 | 0.426 | 0.720 |
| >=80 | 10 | NPV | 0.126 | 0.067 | 0.433 |
| >=80 | 10 | PPV | 0.900 | 0.794 | 0.959 |
| >=80 | 10 | Sensitivity | 0.651 | 0.565 | 0.753 |
| >=80 | 10 | Specificity | 0.390 | 0.236 | 0.936 |

The clinical model included age, baseline CDR-GS, sex, education, APOE ε4, and self-reported race. The augmented model also included $\log_2$ p-tau217. Results used 20 repetitions of 5-fold cross-validation. AUC CIs and differences used 1,000 bootstrap resamples. Brier scores used inverse probability of censoring weighting. Observed calibration risks were Kaplan-Meier estimates within tertiles for participants younger than 60 years and quintiles for the other age groups. Threshold metrics used 0.546 pg/mL and censoring-adjusted risks with 1,000 bootstrap resamples.

Abbreviations: AUC, area under the curve; CDR-GS, Clinical Dementia Rating global score; CI, confidence interval; NPV, negative predictive value; PPV, positive predictive value.
